# Lipidomics and Dietary Assessment of Infant Formulas Reveal High Intakes of Major Cholesterol Oxidative Product (7-ketocholesterol)

**DOI:** 10.1101/2020.11.18.20233528

**Authors:** Alice Kilvington, Carlo Barnaba, Surender Rajasekaran, Mara L. Leimanis, Ilce Gabriela Medina-Meza

## Abstract

Approximately two-thirds of US infants receive infant formula (IF) as a primary or sole nutritional source during the first six months of life. IF is available in a variety of commercial presentations, although from a manufacturing standpoint, they can be categorized in powder-(PIF) or liquid-(LIF) based formulations. Herein, thirty commercial IFs were analyzed in their oxidative and non-oxidative lipidomics profiles. Results show that LIFs have a characteristic lipidomic fingerprint, enriched in an oxidated form of cholesterol, and a lower load of phytosterols. We identified 7-ketocholesterol – a major end-product of cholesterol oxidation – as a potential biomarker of IF manufacturing. Our data allowed re-classification of IF based on their metabolomic fingerprint, resulting in three groups assigned with low-to-high oxidative status. Finally, we modeled the dietary intake for cholesterol, sterols, and 7-ketocholesterol in the first year of life. The database provided in this study will be instrumental for scientists interested in infant nutrition, to establish bases for epidemiological studies aimed to find connections between nutrition and diet-associated diseases, such as sitosterolemia.

## Introduction

Today, several types of infant formula (IF) are available to meet the specific nutritional demands of infants. Soy formulas, hypoallergenic formulas, enriched formulas, *hungry* formulas, goat milk formulas, *good night* milk are some examples of the wide variety of commercial IF presentations. In the United States (U.S.), the main categories of IF are cow’s milk-based, soy-based (for galactosemia), gentle/lactose-reduced (for gastrointestinal issues), and specialty (for more specific issues like phenylketonuria, fat malabsorption, or pre-term) (Maldonado, Gil, Narbona, & Molina, 1998; Martin, Ling, & Blackburn, 2016; Rossen, Simon, & Herrick, 2016; Traves, 2015). The U.S. Special Supplemental Nutrition Program for Women, Infants, and Children (WIC) provides IF for 50% of all US infants and 87% of infants on WIC (Choi, Ludwig, Andreyeva, & Harris, 2020). Approximately 75% of infants in the US receive IF as primary or sole nutrition source during the first six months of life (Harris & Pomeranz, 2020). IF is the gold standard of breastmilk substitutes for infants, and it must contain all the essential nutrients to supply infant needs.

We know from a recent review, that infant metabolites in a healthy population differ based on nutritional intake (M. L. L. Laurens, Kraus-Friedberg, C., Kar, W., Sanfilippo, D., Rajasekaran, S., Comstock, S. S., 2020). Premature infants or sick infants in the intensive care units of hospitals are often fed using IF (M. L. L. Laurens et al., 2020). These patients have weakened immunity from the absence of human milk (Garofalo, 2010; Hassiotou et al., 2013; Horta, Victora, & Organization, 2013; Howie, Forsyth, Ogston, Clark, & Florey, 1990); a developed digestive system and are vulnerable to dietary complications (Santana et al., 2017), due to lower gastric pH (Ouellet, Bailey, & Samson, 2015), or in cases where steroids are used (Group., 1991; Lacroix, Infanterivard, Gauthier, Rousseau, & Vandoesburg, 1986).

Generally, IF is available in two presentations: liquid and powder. Manufacturing of powdered IF (PIF) includes thermal treatments, evaporation, emulsification, pasteurization, sterilization, and spray drying (Gil & Valverde, 1985) to ensure microbial safety and shelf-life. However, exposure to high temperatures (from 170°C to 250°C during processing), light, metal ions, and shear stress among others promotes oxidation reactions, inducing undesirable changes in the product quality (Jiang & Guo, 2014; Kilvington, Maldonado-Pereira, Torres-Palacios, & Medina-Meza, 2019). PIF is usually stored in aluminum cans and vacuum sealed with nitrogen until the consumer first opens it. Liquid IF (LIF) can be either concentrated (needs to be mixed with water before consumption) or ready-to-feed, where sterilization is with high temperatures, followed by aseptic packaging (to guarantee microbial safety). Given its manufacturing, IF is nowadays classified as an Ultra-processed food (UPF) according to the NOVA classification (not acronyms, a name *per se*), which classifies food products according to their extent and purpose of industrial processing (Monteiro et al., 2019; Steele et al., 2016).

In IF approximately 45-50% of the calories come from fat, 8-12% from protein, and 40-45% from carbohydrates (Joeckel & Phillips, 2009). Several bioactive molecules are contained in the lipid fraction such as fatty acid, liposoluble vitamins, and phytosterols (PS). Typically plant-based oils are the major source of fat in IF, mixtures of soy, coconut, palm, safflower, or sunflower are common, being β-sitosterol and campesterol the most common PS found in IF (Hageman, Danielsen, Nieuwenhuizen, Feitsma, & Dalsgaard, 2019; J. Wang, Bertholet, Ducret, & Fleith, 2000). Because of this, IF contains higher amounts of PS than cholesterol. A recent study has shown that both sitosterol and 7β-hydroxysitosterol can cross the blood-brain barrier (Schött et al., 2015). Bovine milk is the most common source of cholesterol in IF with an average of 15-50 mg/L (Babawale et al., 2018). Cholesterol is essential especially during the rapid growth of infants. In addition, fatty acids are other important bioactive lipids, which are essential to forming the brain membrane and myelin (Georgieff & Rao, 2001). Particularly, docosahexaenoic acid (DHA) and arachidonic acid (AA) have been shown to be critical for cognitive, health and development during gestation and early postnatal life; as a result, DHA and AA are often supplemented in IF (Lorenzo et al., 2019).

Lipid oxidation is a natural process that occurs in presence of initiators which may determine the extent of the oxidative process. Usually, malondialdehyde (MDA) is one of the major auto-oxidation products of lipids and so it is commonly used as a biomarker of lipid peroxidation (Botsoglou et al., 1994; Cesa, 2004; Nielsen, Olsen, Jensen, & Skibsted, 1996). Lipid oxidized products have been shown to be harmful to human health (Pozzo et al., 2019; Zmyslowski & Szterk, 2019). MDA, the product of fatty acid oxidation, is suspected to be a carcinogen and lead to mutations (Abela et al., 2019; Griffiths et al., 2016; Kulig, Cwiklik, Jurkiewicz, Rog, & Vattulainen, 2016).

Addition of polyunsaturated fatty acids (PUFA), iron for IF fortification, and antioxidants increase the sensitivity to oxidation, allowing the generation of oxidized compounds such as cholesterol oxidation products (COPs) (Calderon-Santiago, Peralbo-Molina, Priego-Capote, & de Castro, 2012; Maldonado-Pereira, Schweiss, Barnaba, & Medina-Meza, 2018; Medina-Meza & Barnaba, 2013), and MDA derivatives (Cesa, 2004). COPs have been linked to atherosclerosis, hypertension, ischemic stroke, neurological diseases, viral infections, and cancer (Testa, Rossin, Poli, Biasi, & Leonarduzzi, 2018; Willinger, 2019; Zmyslowski & Szterk, 2019). In IF, the evaluation of lipid peroxidation and exposure to oxidized compounds have been focused mainly on adults’ diet; however, IF as UPF may possess a larger number of oxidized compounds and the adverse effects on an infant’s health is still unknown. Considering health risks associated with oxidized lipids intake, the oxidative load on IF should be studied.

In this study, we performed a large-scale study on the oxidative status of IF. Thirty commercial products were collected and their lipidomic profiles were assessed, with particular attention to oxidized cholesterols and oxidative markers. To the best of the authors’ knowledge, ours is the first study that provides a vast lipidomic analysis in a large set of commercial formulas. We confirm that 7-ketocholesterol is a biomarker for formula manufacturing and a potential biomarker of IF consumption through dietary exposure model. Based on our lipidomic dataset, a dietary intake assessment was performed for the 7-ketocholesterol biomarker, as well as overall oxidative load. Finally, we regroup the formulas according to their oxidative status using a clustering algorithm, overcoming the limitation of labeling attributes with a more rigorous metabolomic-based classification approach. The presented results will serve as a guide and data reference for consumers, pediatricians, clinicians, and scientists interested in infant diet and development.

## Materials and Methods

### Chemicals

Diethyl ether, ethanol, chloroform, methanol, hexane, ethyl acetate, isopropyl alcohol, acetone, sulfuric acid, sodium sulfate, potassium hydroxide, sodium chloride, campesterol, tocopherols, stigmasterol, squalene, β-sitosterol, and Supelco 37 FAME mix were purchased from Sigma Aldrich (St Louis, MO). Cholesterol, fucosterol, brassicasterol, lanosterol, 5α-cholestane, 19-hydroxycholesterol, and the cholesterol oxidation products (COPs) standards were purchased from Steraloids Inc. (Newport, Rhode Island).

### Infant formulas sample set

A total of thirty IF both powder and liquid were purchased from local supermarkets and/or online through the manufacturer’s website, covering a broad variety of nutrition requirements for infants, and commonly used in a hospital setting. The nutritional contents of the IF as indicated in the label is presented in **Table S1**.

### Lipid extraction

Lipids were extracted using the Folch method (Folch, Lees, & Sloane-Stanley, 1957) with a few modifications. Briefly, IF powders were solubilized in water according to the directions on the label. One scoop (9 grams) of powder was mixed with 30 mL of tap water to emulate the common preparation for consumption. The extraction of the lipid fraction was done with 8:1 solution of diethyl ether: ethanol. After phase separation, lipid fraction was recovered, dried under N_2_ stream, resuspended in 4:1 hexane: isopropyl alcohol, and stored at -20°C for further analysis. A similar procedure was used with the ready-to-eat LIF; however, due to the excessive water being collected in the lipid fraction, methanol was used instead of ethanol and chloroform was used instead of diethyl ether for extraction. We did not observe any difference in fat yield extraction by changing the solvents mixture. LHF3, a concentrated liquid formula, had to be diluted 1:1 with water before the Folch procedure.

### Total sterols profiling by GC-MS

Two hundred mg of fat were placed into a glass tube and dried under a nitrogen stream. 40 ug of 5α-cholestane (5α) and 50 ug of 19-hydroxycholesterol (19-OH) were added as internal standards. Ten mL of 1 N methanolic KOH were added and vortexed. The unsaponifiable fraction was collected and sodium sulfate was added. The mixture was left standing for 2 hours and then filtered over a bed of sodium sulfate and collected in a round bottom flask. The solution was then dried in a vacuum evaporator. The sterols were then collected in diethyl ether and dried with a stream of nitrogen. One mL of 4:1 hexane: isopropanol was added and then stored at -20°C until analysis. One-hundred microliters (µL) were derivatized by drying with nitrogen and adding 100 µL of pyridine and 100 µL of silanization solution and then heated at 60°C for 40 min. The solution was then dried and resuspended in 200 µL hexane. Two µL were injected into a GC coupled to a MS (GC-MS) (Shimadzu GCMS-QP 2010 SE, LabSolutions GCMS Solution Version 4.45). Injector temperature was set at 320°C. The oven temperature profile was programmed to start at 260°C and increase to 300°C at a rate of 2.5°C/min, then from 300°C to 320°C at a rate of 8°C/min and held for 1 min. Helium was used as a carrier gas with a pressure of 134 kPa. The sterol identification was done by comparing retention times and mass fragmentations from pure standards. Quantification was done by comparing peak areas using the internal standards.

### Sterols oxidation products profiling by GC-MS

The remaining 900 µL of the unsaponifiable fat were enriched in the sterol oxidation products by solid-phase extraction (SPE), using NH_2_ Strata® SPE cartridge. Enriched samples were dried using a nitrogen stream and then derivatized as described above. First, the samples were analyzed using the Total Ion Chromatogram (TIC) method. Two µL of the sample were injected into a GC coupled to a MS (GC-MS) (Shimadzu GCMS-QP 2010 SE, LabSolutions GCMS Solution Version 4.45). The injector temperature was set to 320°C. The oven temperature profile was programmed from 260°C to 280°C at a rate of 2°C/min and held for 7 min and then from 280°C to 315°C at a rate of 1.5°C/min. Identification of compounds was performed using retention times and mass fragmentations from commercial standards. Using the TIC method, the quantification of the total sterols content was done by comparing the peak areas to the internal standard, 19-OH. The Selected Ion Monitoring (SIM) method was used to profile the COPs in the IF. The injector temperature was set to 320°C. The oven temperature profile was programmed from 260°C to 280°C at a rate of 2.0°C/min and held for 7 min and then from 280°C to 315°C at a rate of 1.5°C/min. Helium was used as a carrier gas with a pressure of 49.2 kPa and a column flow rate of 0.41 mL/min. The ion source temperature for the MS was set at 230°C. Identification of compounds was done using retention times and mass fragmentations from commercial standards.

### Fatty acid methyl esters profiling (FAME)

FAME samples were prepared according to Chen et al. (Chen, Aluwi, Saunders, Ganjyal, & Medina-Meza, 2019). Twenty milligrams (mg) of fat were placed into a test tube. Tridecanoic acid was used as an internal standard. Briefly, 0.5 N NaOH in methanol was added to the samples, heated for 15 min at 105ºC, and then cooled on ice for one minute. Boron trifluoride was added and samples were heated for 15 min, following by rapid cooling in an ice bath. One mL of hexane and 2 mL of 2M NaCl were added to the samples and then shaken vigorously. The methylated fraction was collected and diluted 200-fold. One µL was injected into a gas chromatograph coupled with flame ionization detector (GC-FID) (Shimadzu GC-2010, Kyoto, Japan). The injector temperature was set at 250ºC. The oven temperature started at 120ºC and increased to 200°C at a rate of 3.5°C/min, then further increased to 240°C at a rate of 1.5 ºC /min and then was held for 2 min. Helium was used as carrier gas with a pressure of 92.2 kPa and a total flow rate of 5.9 mL/min. Identification was done by comparison of retention times of FAME standards (FAME Supelco 37 mixture Sigma Aldrich). A standard curve with 500, 200, 100, 50, and 25 µg/mL concentrations were used for quantification.

### Oxidative status (TBARS)

The thiobarbituric acid reactive substances (TBARS) assay was performed following a modified version of Miller’s protocol (Miller, 1998). Sixty mg of fat from the infant formulas were placed into a test tube. One-hundred µL of butylated hydroxytoluene was added to each sample. One set of replicates was spiked with 1,1,3,3-tetraethoxypropane (TEP) to confirm the reaction was occurring. Then, 10% tricholoroacetic acid (TCA) in phosphoric acid solution was added. Samples were mixed and then split into two aliquots. One aliquot was mixed with deionized water to be used as a blank and the other aliquot was mixed with thiobarbituric acid (TBA). Samples were left overnight in a dark cabinet. Standards of 0.5, 1.0, 1.5, 2.0 mL TEP were mixed with TCA and TBA and leftover night to be used as a calibration curve. Absorbance was recorded with a UV–Vis Double Beam Spectrophotometer UV-6300PC (VWR, Radnor, PA) at 530 nm. Quantification was achieved by comparing the sample absorbances with the calibration curve, and results expressed as mg of MDA/g fat.

### Total antioxidant capacity (ToAC)

The ToAC was performed according to Khan and co-workers (Khan et al., 2017). Briefly, 3 mL of 0.6 M sulfuric acid, 3 mL of 4 mM ammonium molybdate, and 3 mL of 28 mM sodium phosphate was added to 0.3 mL of IF. The test tubes were incubated at 85ºC for 90 min. After cooling to room temperature, the mixture was centrifuged, and the supernatant was placed into a glass cuvette and read using a UV–Vis Double Beam Spectrophotometer UV-6300PC (VWR, Radnor, PA) at 695 nm. Results were reported as mg equivalents of ascorbic acid/L.

### Protein

Two-hundred and fifty mg of IF was used to determine the protein content by using a LECO Nitrogen Analyzer FP828. The method used was provided by LECO with coordination from the Application Specialist to be optimized for the IF matrix. EDTA was used as a standard to calibrate the machine, and helium was used as a carrier gas. The furnace was set to 950°C and the afterburner was set at 850°C. The furnace flow was set to 5 L/min for 30 seconds, then 1 L/min for 30 seconds, and then 5 L/min. The standard protein factor of 6.38 was used to obtain protein content.

### Dietary intake

A dietary intake (DI) assessment spanning the first 12 months of infant life was performed. The DI for the first six months was calculated based on the energetic requirements (Food and Agriculture Organization, 2004), assuming a 100% formula diet. At 6 months, infants are introduced to solid food according to USDA guidelines, thus from 6 to 12 months an average IF supplement to a solid diet was used (USDA, 2018). 7-ketocholesterol, Total COPs, Total Phytosterols, and Total Sterols intakes were assessed using mean and the 95% confidence interval (**Table 1**). The model used for calculating the DI was the following:

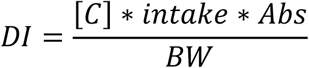

**Table 1.** Content of fat, steroidal compounds, fatty acids, and redox values for in IF. Data are expressed as mg/serving except where indicated. Average and the 95% confidence interval is reported for the whole dataset (IF, *n*=30) and powder (PIF, *n*=15) and liquid (LIF *n*=15) categories. *t-*test significance assessment between PIF and LIF is indicated. ^¶^ Sum of steroids from animal origin. ^ǂ^ Sum of phytosterols.

| | IF | PIF | LIF | $t$-test (PIF vs. LIF) |
| --- | --- | --- | --- | --- |
| Protein (g/serving) | 1.9 (1.7-2.1) | 1.6 (1.5-1.7) | 2.2 (1.7-2.6) | *** |
| Total fat (g/serving) | 2.57 (2.41-2.74) | 2.71 (2.47-2.96) | 2.43 (2.20-2.66) |  |
| *Sterols* |  |  |  |  |
| Cholesterol | 1.60 (1.11-2.08) | 1.34 (0.57-2.11) | 1.94 (1.14-2.74) |  |
| Desmosterol | 0.23 (0.21-0.26) | 0.26 (0.21-0.31) | 0.21 (0.18-0.23) |  |
| Stigmasterol | 0.13 (0.11-0.14) | 0.14 (0.12-0.16) | 0.11 (0.01-0.13) | * |
| β-sitosterol | 0.67 (0.59-0.76) | 0.81 (0.71-0.92) | 0.53 (0.43-0.63) | *** |
| Campesterol | 0.46 (0.38-0.55) | 0.60 (0.48-0.72) | 0.33 (0.27-0.38) | *** |
| Fucosterol | 0.14 (0.11-0.17) | 0.18 (0.14-0.22) | 0.11 (0.08-0.13) | ** |
| Total Sterols<sup>¶</sup> | 1.87 (1.33-2.40) | 1.60 (0.83-2.37) | 2.13 (1.33-2.93) |  |
| Total Phytosterols<sup>‡</sup> | 2.64 (2.24-3.04) | 3.33 (2.77-3.89) | 1.95 (1.64-2.26) | *** |
| *Tocopherols* |  |  |  |  |
| δ-tocopherol | 0.15 (0.10-0.19) | 0.20 (0.12-0.29) | 0.09 (0.06-0.12) | * |
| β-tocopherol | 0.019 (0.011-0.027) | 0.014 (0.008-0.021) | 0.024 (0.009-0.039) | |
| γ-tocopherol | 0.019 (0.011-0.027) | 0.033 (0.023-0.043) | 0.006 (0.002-0.013) | *** |
| α-tocopherol | 0.044 (0.031-0.057) | 0.032 (0.020-0.043) | 0.056 (0.033-0.078) | |
| Total Tocopherols | 0.22 (0.17-0.28) | 0.28 (0.18-0.38) | 0.17 (0.11-0.22) | * |
| Squalene | 0.059 (0.036-0.082) | 0.060 (0.025-0.10) | 0.060 (0.0240.092) |  |
| *COPs (μg/serving)* |  |  |  |  |
| 7α-hydroxycholesterol (7α-OH) | 5.20 (3.68-6.71) | 4.22 (2.67-5.78) | 6.17 (3.43-8.89) | |
| 7β-hydroxycholesterol (7β-OH) | 6.75 (4.54-8.95) | 5.24 (2.70-7.78) | 8.25 (4.50-12.02) | |
| 7-ketocholesterol (7-keto) | 1.42 (0.94-1.89) | 1.97 (1.20-2.75) | 0.86 (0.39-1.33) | ** |
| 25-hydroxycholesterol (25-OH) | 0.94 (0.31-1.58) | 1.09 (0.51-1.65) | 0.81 (-0.42-2.03) |  |
| 3,5,6-cholestan-triol (triol) | 1.12 (0.79-1.44) | 1.29 (0.70-1.87) | 0.95 (0.62-1.28) |  |
| 6-ketocholestenol (6-keto) | 1.07 (-0.01-2.16) | 1.95 (-0.26-4.15) | 0.20 (0.06-0.33) |  |
| 5β,6β-epoxycholesterol (5,6β-epoxy) | 0.21 (0.13-0.29) | 0.22 (0.10-0.34) | 0.20 (0.07-0.33) | |
| 5α,6α-epoxycholesterol (5,6α-epoxy) | 0.17 (0.08-0.27) | 0.24 (0.05-0.44) | 0.11 (0.05-0.16) | |
| Total COPs | 19.2 (12.3-26.1) | 16.8 (10.9-22.7) | 21.6 (8.3-34.9) | * |
| *FAME (%)* |  |  |  |  |
| C8 | 8.6 (5.6-11.7) | 3.3 (0.8-5.9) | 6.9 (3.2- 10.5) |  |
| C10 | 5.6 (3.9-7.2) | 2.7 (0.8- 4.5) | 5.6 (2.7- 8.6) |  |
| C12 | 6.9 (5.7- 8.1) | 8.8 (7.4-10.2) | 5.2 (2.8-7.6) | ** |
| C14 | 3.4 (2.7- 4.1) | 4.0 (3.3- 4.6) | 2.5 (1.3-3.6) | * |
| C16 | 14.4 (12.1-16.6) | 15.5 (11.3-19.8) | 12.7 (8.4-16.9) |  |
| C16:1 | 0.07 (0.06-0.09) | 0.10 (0.07-0.13) | 0.07 (0.04-0.09) |  |
| C18 | 3.9 (3.6- 4.2) | 3.7 (3.3- 4.2) | 4.7 (4.1-5.4) |  |
| C18:1cis | 32.0 (28.8-35.1) | 36.9 (34.6-39.1) | 39.3 (37.6-41.0) |  |
| C18:2cis | 21.4 (19.7-23.2) | 21.2 (19.7-22.7) | 19.5 (18.2-20.8) |  |
| C18:3n6 | 2.3 (2.1-2.5) | 2.3 (2.1- 2.5) | 2.0 (1.8-2.2) |  |
| C20 | 0.25 (0.22-0.27) | 0.29 (0.27-0.31) | 0.31 (0.27-0.34) |  |
| C20:1 | 0.11 (0.09-0.13) | 0.13 (0.12-0.15) | 0.15 (0.11-0.19) |  |
| C20:3n6 | 0.59 (0.52-0.66) | 0.48 (0.41-0.55) | 0.55 (0.45-0.64) |  |
| C20:5 | 0.21 (0.18-0.25) | 0.18 (0.15-0.21) | 0.22 (0.15-0.30) |  |
| C24 | 0.23 (0.20-0.27) | 0.20 (0.15-0.26) | 0.20 (0.15-0.24) |  |
| SFA (%) | 43.3 (39.1-47.4) | 38.3 (36.0-40.7) | 48.2 (40.7-55.8) | * |
| MUFA (%) | 32.1 (27.6-36.7) | 38.3 (36.3-40.4) | 26.0 (17.8-34.1) | ** |
| PUFA (%) | 24.6 (21.9-27.2) | 23.3 (21.8-24.8) | 25.8 (20.5-31.1) |  |
| TBARs (ppb/serving) | 0.52 (0.49-0.55) | 0.50 (0.49-0.52) | 0.53 (0.47-0.59) |  |
| ToAC (mg ascorbic ac. equiv./serving) | 7.4 (4.3-10.6) | 10.7 (5.0-16.4) | 4.2 (1.8-6.6) | * |

COPs: Cholesterol oxidation products, TBARs: Thiobarbituric acid reactive substances, ToAc: Total Antioxidant (equivalent Ascorbic acid). SFA: saturated fatty acid, MUFA: monounsaturated fatty acid, PUFA: Polyunsaturated fatty acid.

Where DI is the dietary intake (mg kg^-1^ of body weight d^-1^), *[C]* is the concentration of the compounds (mg mL^-1^ of formula, as liquid or prepared from powder), *intake* is the average daily formula intake during the specific growth period (mL d^-1^), *Abs* is the coefficient of absorption of the considered compounds, and *BW* is the average body weight of infants during the specific growth period (kg). There is no consensus on the *Abs* percentage of COPs and sterols. For the parent cholesterol, ranges from 20 to 80% have been published (Ogino et al., 2007; Vine, Croft, Beilin, & Mamo, 1997), with 60-70% being the most reported. For phytosterols, higher absorption than cholesterol has been reported (Grandgirard, Sergiel, Nour, Demaison-Meloche, & Gihiès, 1999; Liang et al., 2011; M. M. Wang & Lu, 2018), although results are controversial. Herein, we opted for a conservative 0.5 coefficient. The average *BW* for infants was obtained from the literature (De Onis & Onyango, 2008); 50^th^ percentile of weight was used. We found no appreciable differences between female and male dietary intake; thus, data were averaged between sexes.

### Statistical analysis

A one-way ANOVA (95% significance level) was performed to statistically evaluate the chemical responses across the different formulations; Pearson correlation across variables was also tested. Variables expressed as percentage (i.e. protein) were arcsine square root transformed (Olson, 1976). Before variable reduction analysis, the dataset was normalized by sum, mean centered, auto scaled, and the normal distribution verified (Wanichthanarak, Fan, Grapov, Barupal, & Fiehn, 2017). ANOVA-based heatmap and biomarker analysis, including the ROC curves, were performed using MetaboAnalyst in its web server version (Chong et al., 2018; Xia & Wishart, 2016). The transformed data were subjected to Factor Analysis to reduce the variables to a set of new orthogonal variables retaining >70% of the variance across the samples. Before Factor Analysis, data adequacy was assessed by the Kaiser-Meyer-Olkin (KMO) test (Kaiser, 1974), and variables having KMO a value <0.5 were excluded and factor analysis repeated until the overall KMO value was 0.6 or higher. Factor analysis was performed using the Principal Components Analysis algorithm. Identification of the minima number of PCs was achieved by cumulative variance and direct visualization of the scree plot (Cadima & Jolliffe, 1995). The first five components, representing ∼73% of the cumulative variance, were used for subsequent cluster analysis. The *k-*means clustering algorithm was performed using the hierarchical Ward’s criterion (minimum variance method); distance between clusters was computed using the Euclidean algorithm. PCA’s eigenvalues were visualized using a heatmap algorithm. ANOVA, Pearson correlation, KMO test were computed using SPSS (IBM). PCA and *k-means* clustering were performed in OriginPro (OriginLab, v.2020).

## RESULTS

### Overall metabolite and oxidative status assessment

We performed fat and protein quantification to determine if our findings matched the information contained in the product label (**Table S2** in S*upplemental information*). The total fat recovered after extraction from the LIF was much closer to the label compared to PIF (14% vs. 28% difference on average). LIFs have significant lower protein content than PIF (*p* < 0.01). Compared to the amount reported in the label, our results underestimated the protein content on an average of 27%; the difference between the reported value and our findings were 36% and 18% for PIF and LIF, respectively. The correlation between label and measured protein content was high in PIF (*R* = 0.93), but poor in LIF (*R*^*2*^ = 0.34). The reason for such difference between label and measured data can be attributed to several factors, including the method and/or equipment used for quantifying protein. The liquid formulations used in this study came in different presentation (ready-to-fed, concentrated), thus the preparation of sample prior to analysis can be an additional source of analytical error. When compared to literature, the results for both fat and protein align (Koletzko, 2009; Lorenzo et al., 2019; Saxena et al., 2019).

Lipidomics showed a higher amount of total phytosterols in the PIF than the LIFs, with an average of 3.33 (2.77-3.89, 95% CI) mg/scoop and 1.95 (1.64-2.26) mg/scoop, respectively. Cholesterol was found with an average content 1.60 (1.11-2.08) mg/scoop, followed by β-sitosterol with 0.67 (0.59-0.76) mg/scoop, campesterol 0.46 (0.38-0.55) mg/scoop, desmosterol 0.23 (0.21-0.26)mg/scoop, fucosterol 0.14 (0.11-0.17) mg/scoop, and stigmasterol 0.13 (0.11-0.14) mg/scoop. Comparison with previous literature is difficult, due to the lack of large datasets, and the units of quantitative measurement. Two studies on IF estimated the total sterol concentration to be in the range of 6.5-15.2 mg/100 mL reconstituted IF (G. García-Llatas et al., 2008; Hamdan, Sanchez-Siles, Garcia-Llatas, & Lagarda, 2018). Comparatively, we found total sterol concentrations of 6.46 ± 1.64 mg/100 mL of sample and 11.08 ± 2.11 mg/100 mL of IF liquid and powder, respectively. In contrast, cholesterol concentrations in IF may vary between 0.1 to 7.3 mg/g lipid (Scopesi et al., 2002; Zunin, Calcagno, & Evangelisti, 1998). Our results range from 0.3 to 2.14 mg of cholesterol per g of lipid (**Table S3**, *Supplemental information*).

Regarding bioactive lipids with antioxidant properties, we found tocopherols. α-, β-, γ-, and δ-tocopherols belong to the vitamin E class of compounds. These compounds are known for their radical scavenging properties, for protecting against chromosome damage, lipid peroxidation, and DNA oxidation (Li, Cherian, Ahn, Hardin, & Sim, 1996). Tocopherols are usually added to IF to increase vitamin content and prevent lipid oxidation during processing and storage, extending the product’s shelf-life (Miquel, Alegria, Barbera, Farre, & Clemente, 2004). Total tocopherols range from 0.07 to 0.71 mg/scoop in PIF and from 0.054 to 0.33 mg/serving in LIF (**Table S4** *Supplemental material*). Overall, PIF contained higher amounts of tocopherol compared to LIF (*p* < 0.05, **Table 1**), with the γ-tocopherol being 5-fold higher in powder formulations. These results agree with Lee et al. (Lee, Jang, & Kim, 2013) and Miquel et al (Miquel et al., 2004) for powder IF and contrast with Dorota et al. (Martysiak-Zurowska, Szlagatys-Sidorkiewicz, & Zagierski, 2013) for liquid IF. These differences may be accounted to the type of formula used (a specific *follow-up* IF), as well as manufacturing. Little information has been reported for COPs and their metabolites in IF (Scopesi et al., 2002). 7α-hydroxycholesterol (7α-OH), 7β-hydroxycholesterol (7β-OH), 7-ketocholesterol (7-keto), 5α,6α-epoxycholesterol (5,6α-epoxy), 5β,6β-epoxycholesterol (5,6β-epoxy), cholestane-3β,5α,6β-triol (triol), and 25-hydroxycholesterol (25-OH) (Gorassini, Verardo, Fregolent, & Bortolomeazzi, 2017; Guardiola, Bou, Boatella, & Codony, 2004; Rodriguez-Estrada, Garcia-Llatas, & Lagarda, 2014) and 7-ketocholesterol are the most common COPs found in foods (Zunin et al., 1998).

We found an average of 16.8 (10.9-22.7, 95% CI) μg/scoop and 21.6 (8.3-34.9, 95% CI) μg/scoop for powder and liquid formulations, respectively, is significantly higher for LIF (*p*<0.05). Nine COPs (7β-OH, 7α-OH, 7-keto, 22-OH, 25-OH, triol, 6-keto, 5,6β-epoxy, and 5,6α-epoxy) were identified in the IF samples. The most abundant were the C7-hydroxy isomers 7β-OH (average of 6.75 µg/scoop) and 7α-OH (average of 5.20 µg/scoop). Garcia-Llatas et al. (G. García-Llatas et al., 2008) reported COPs found in two ready-to-eat IFs (23-28 µg/100 g sample), which is lower than our data when adjusted to the amount of sample. We cannot exclude that this discrepancy could be due to differences in the manufacturing process between Europe and the USA (Guo, 2014). 7-keto was the third highest COPs in IF, with an average value of 1.42 μg/scoop (0.77μg/g lipids for PFI and 0.49 μg/g lipids for LIF); our results agree with Scopessi and coworkers (Scopesi et al., 2002) who found 0.3 – 11.2 µg/g of lipid of 7-keto from a set of ten IFs. Conversely, our data is lower than that reported by others (1.2 – 11.2 µg/g of lipids), in a study performed on six IF (Zunin et al., 1998). It is not possible to exclude that such discrepancies in COPs and other oxidative compounds between studies can be associated with the advancement in IF manufacturing process and formulation itself. Importantly, our data show that 7-keto is the only COPs present in significantly (*p*<0.01) higher amount in PIF, compared to LIF.

Saturated fatty acids (SFA) made up 43 ± 11% of the fatty acids, with C16:0 as the most abundant fatty acid in all IF, followed by C8:0 and C:10:0. These results are in line with previous studies, confirming that SFA% may be a good biomarker for milk-based fat IF (Ceballos et al., 2009). Comparing formulations, LIF contained up to 2-fold higher amount of C:8 and C:10 than PIF; however, the differences were not statistically significant (*p*=0.10 and *p*=0.08, respectively). On the contrary, C12 and C14 were slightly but significantly higher in PIF (**Table 1**). Monounsaturated fatty acid (MUFA) average content is 32 ± 12% with C-18:1 being the most abundant fatty acid. PIF contained ∼45% more MUFA than LIF (*p* < 0.01), at the expenses of SFA content. It is known that oleic acid content is higher in plant-based oil source IF, due to their high content in sunflower and soybean oils. Polyunsaturated fatty acid (PUFA) content was 25 ± 7%, with C18:2 as the major fatty acid followed by C:18:3n6. No significant differences were found between LIF and PIF. Many other research groups have analyzed the fatty acid profile of IF, being close to our findings (Guadalupe García-Llatas et al., 2008; Hageman et al., 2019; Romeu-Nadal, Chávez-Servín, Castellote, Rivero, & López-Sabater, 2007). The major difference in FAME profile is the oil source, which plays a critical role in the formulation, plant-oil formula and cow’s milk formula are the most common formulations followed by goat’s milk IF (Sun et al., 2016).

TBARS and ToAC values are also difficult to compare with literature, given the measurement units adopted. For TBARs, we found an average of 0.52 ppb (0.49-0.55 ppb, 95% CI), which is comparable only with the amount found by Turoli et al. (Turoli, Testolin, Zanini, & Bellù, 2004) and Michaski et al. (Michalski, Calzada, Makino, Michaud, & Guichardant, 2008). Other reports have inconsistently high values >100 ppb (Cesa, 2004). ToAC, measured as ascorbic acid equivalents, is a direct estimation of the overall antioxidant capacity (Boatright & Crum, 2016). In our dataset, we have an average of 7.4 mg/mL (4.3-10.6 mg/mL); however, LIF had roughly double the amount that PIF.

### Load of steroids and cholesterol oxidative products in Ifs

Several non-oxidative and oxidative markers characterize the powder formulation. The Volcano plot for the lipidomics markers quantified in the present study is depicted in **Figure 1**. Among non-oxidative markers, PIF has higher amounts of phytosterols, including tocopherols, fucosterol, and campesterol. Tocopherols are supplemented in formulas (Martysiak-Zurowska et al., 2013; Miquel et al., 2004), or derived from the vegetable oils used for their formulation (Manglano et al., 2005) for increasing the antioxidant capacity and prevent lipid peroxidation and thus reduce oxidative stress (Jialal, Devaraj, & Venugopal, 2002). Phytosterols also are derived from the vegetable oils used for formula preparation. Some phytosterols are naturally present in breast milk, as campesterol, while fucosterol is not usually found in human milk (Hamdan et al., 2018). Among oxidative markers, the powder IF has several folds larger amounts of individual COPs (7-keto, triol, and 25-OH), as well as MDA content. It is worth to note that 7-keto is an end-product of cholesterol oxidation and is derived from further oxidation of 7-hydroxyl epimers. Similarly, triol is the end-product of 5,6-epoxides, and 25-OH is the most important derivative of cholesterol oxidation in its side chain (Medina-Meza, Rodriguez-Estrada, Garcia, & Lercker, 2012; Medina-Meza, Rodríguez-Estrada, García, & Lercker, 2012; Medina-Meza, Rodriguez-Estrada, Lercker, Barnaba, & García, 2014) The ANOVA-based heatmap in **Figure 1B** is helpful to identify characteristics of abundance that are predominant in the powder vs. liquid presentation. At the same time, **Figure 1B** suggests a potential subclassification based on a more specific lipid and oxidative signature. Strikingly, a subpopulation of PIFs showed higher content in several COPs, which potentially masks the fact that only 7-keto and triol are spiking in the Volcano plot.

**Figure 1.**
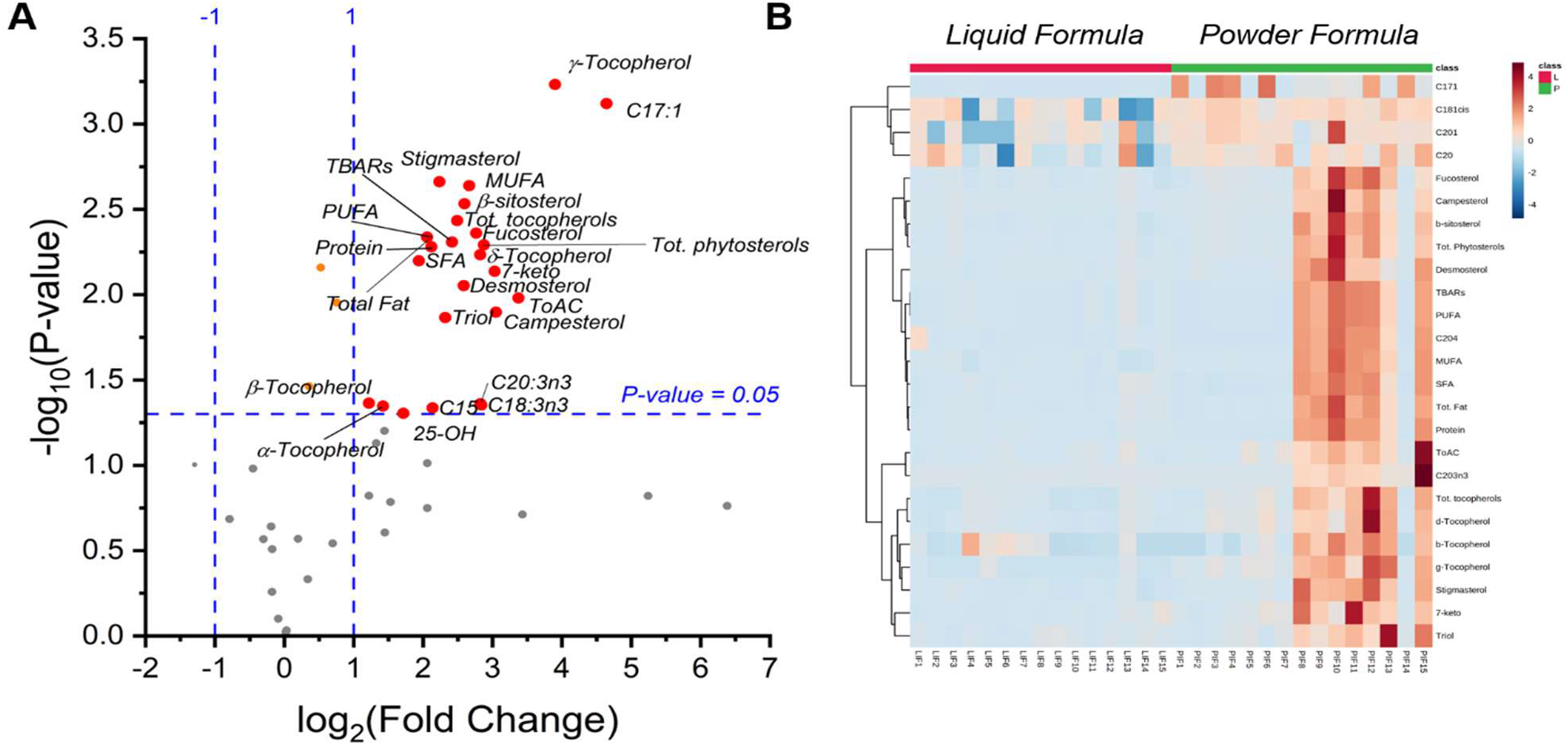
Metabolomic signature of powder (PIF) vs liquid (LIF) infant formulas. **A)** Volcano plot showing significant contrast between PIF and LIF. Compounds or classes of compounds with >2-fold higher amount per serving in PIF at p=0.05 are depicted with red dots. **B)** ANOVA heatmap, showing significant differences between IF (p<0.05).

### Biomarker of the oxidative process

Next, we performed a ROC analysis to identify potential biomarkers as a signature of formula manufacturing. γ-tocopherol followed by campesterol gave higher AUC values (>0.95, *p*<0.001); however, they are not indicative of the process *per se*, but the ingredients used for formulation. On the other hand, COPs and the oxidative load are a direct consequence of the food process history (Savage, Dutta, & Rodriguez-Estrada, 2002; Tamanna & Mahmood, 2015). In the biomarker analysis, 7-keto showed good discriminant potential (AUC=0.86, *p*<0.01) **(Fig. 2A,B)**, which was also evident from the *t*-test **(Table 1)**. For instance, 7-keto is a direct derivative of cholesterol autooxidation, which is exacerbated during food processing, particularly those operations that include heat, as thermal sterilization (Rodriguez-Estrada et al., 2014; Zardetto, Barbanti, & Rosa, 2014).

**Figure 2.**
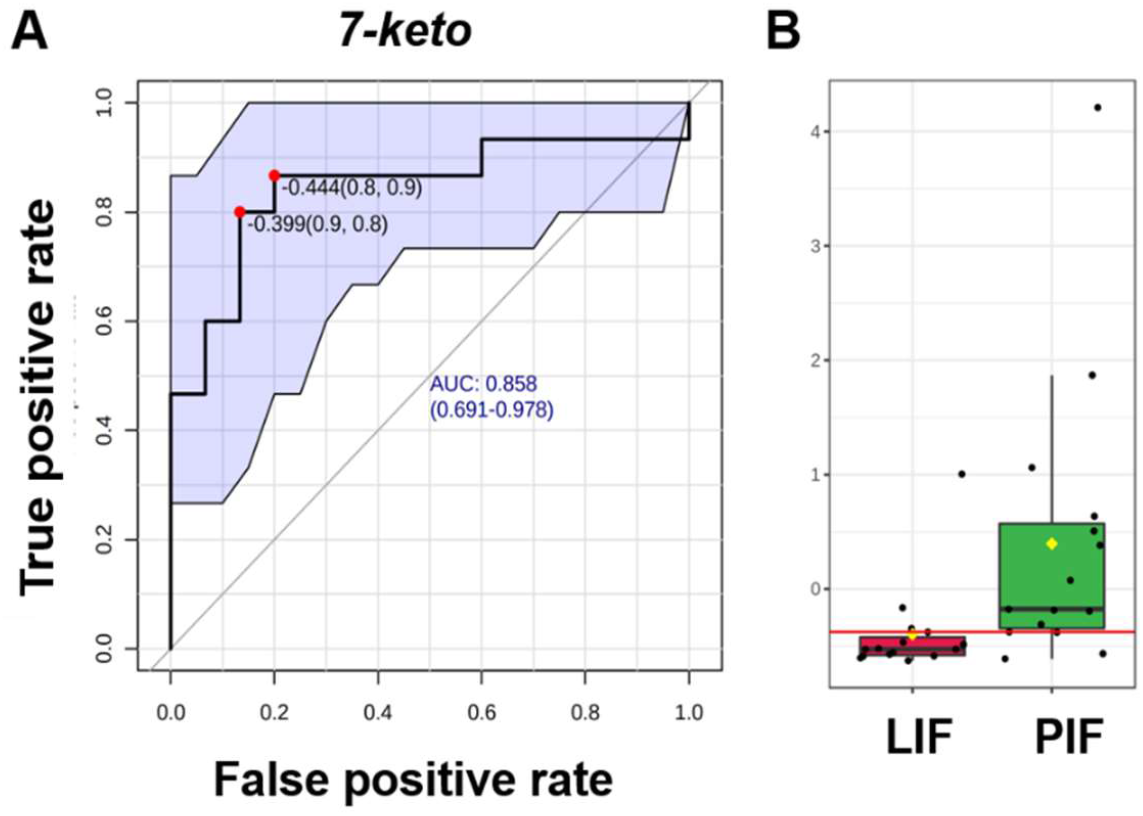
7-keto as potential biomarker for IF processing. **A)** ROC curve for 7-keto. **B)** Boxplot of 7-keto. The notch indicates 95% CI around the median of each group. The yellow diamond indicates the mean concentration for each group.

### Reclassification of infant formulas based on metabolite fingerprint

The heatmap presented in Fig. 1B hints at a subclassification based on metabolite fingerprint rather than commercial presentation. Thus, we performed a PCA-based factor analysis followed by hierarchical classification to re-group the IF based on selected variables explaining the majority of variance across metabolic responses. PCA reduces the dimensionality of the data set; each principal component represents a new variable in which the original metabolite responses have a “weight” (**Figure 3A**). The first three principal components retained >60% variance and were retained for further statistical assessment **(Figure 3A)**. Based on the score of each IF, three groups were identified (**Figure 3B):** *i*) IFs having highest COPs content, with low phytosterols and tocopherols; this group is also characterized for having low ToAC. Members of this group (green in the dendrogram) are equally distributed among PIF and LIF; *ii)* IFs having higher PC1 scores, corresponding with high phytosterols amount and moderate-to-high oxidative load (COPs), including the cytotoxic 5,6-epoxides epimers, 6-keto, and 7-keto. This group (red in **Figure 3B)** consists of the great majority of LIF formulas (7 of 8 total). Finally, *iii)* IFs with the lowest load in autooxidation products of cholesterol. The third group (blue in the dendrogram) is more heterogeneous, it mainly contains PIF.

**Figure 3.**
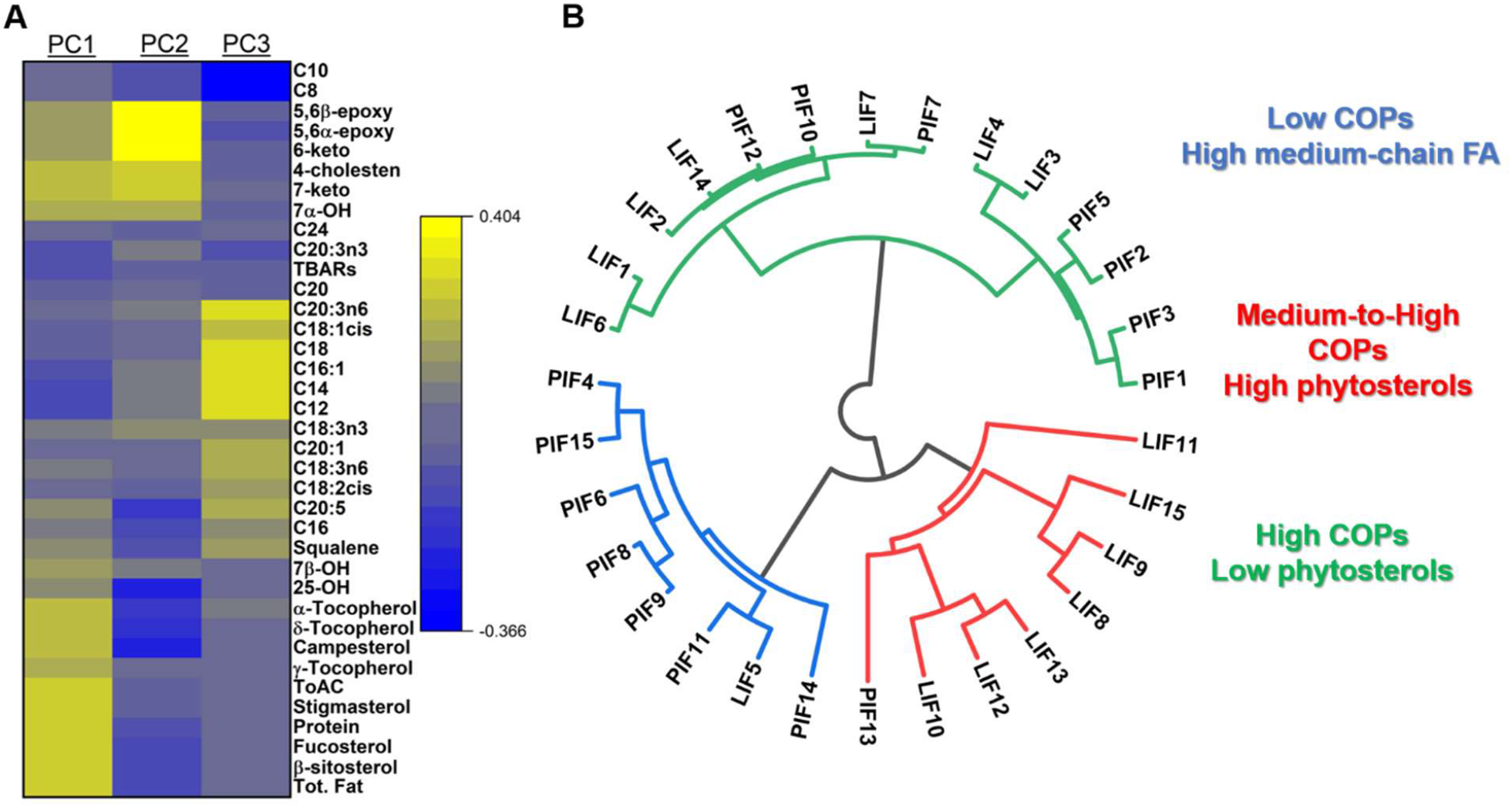
Re-classification of IFs using multivariate data analysis and hierarchical clustering. **A)** Heat-map representing the metabolomics variables and their scores associated with the PCs. **B)** Dendrogram depicting the classification of IFs according to the PCA variables.

### Dietary intake of 7-keto, COPs, sterols, and phytosterols

Infants consume milk, either breast milk or formulations, as the only food intake for the first 6 months of their life (Harris & Pomeranz, 2020). After 6 months, and coinciding with weaning, milk is integrated with solid food. We estimated the dietary intake for total COPs, total animal sterols, total phytosterols, and our identified biomarker 7-keto (**Fig. 4**) over the first year of infant life. Overall, the daily intake decreases by half once weaning sets in; after six months, the dietary intake does not reflect the exposure to these compounds, since we are not taking into account sterols and oxidative derivatives present in solid foods. Compared to phytosterols, there is higher variability in COPs, sterols, and 7-keto intakes, given the broad distributions in our IF dataset. The dietary intake of phytosterols is roughly double than animal sterols **(Fig. 4B,C)**.

**Figure 4.**
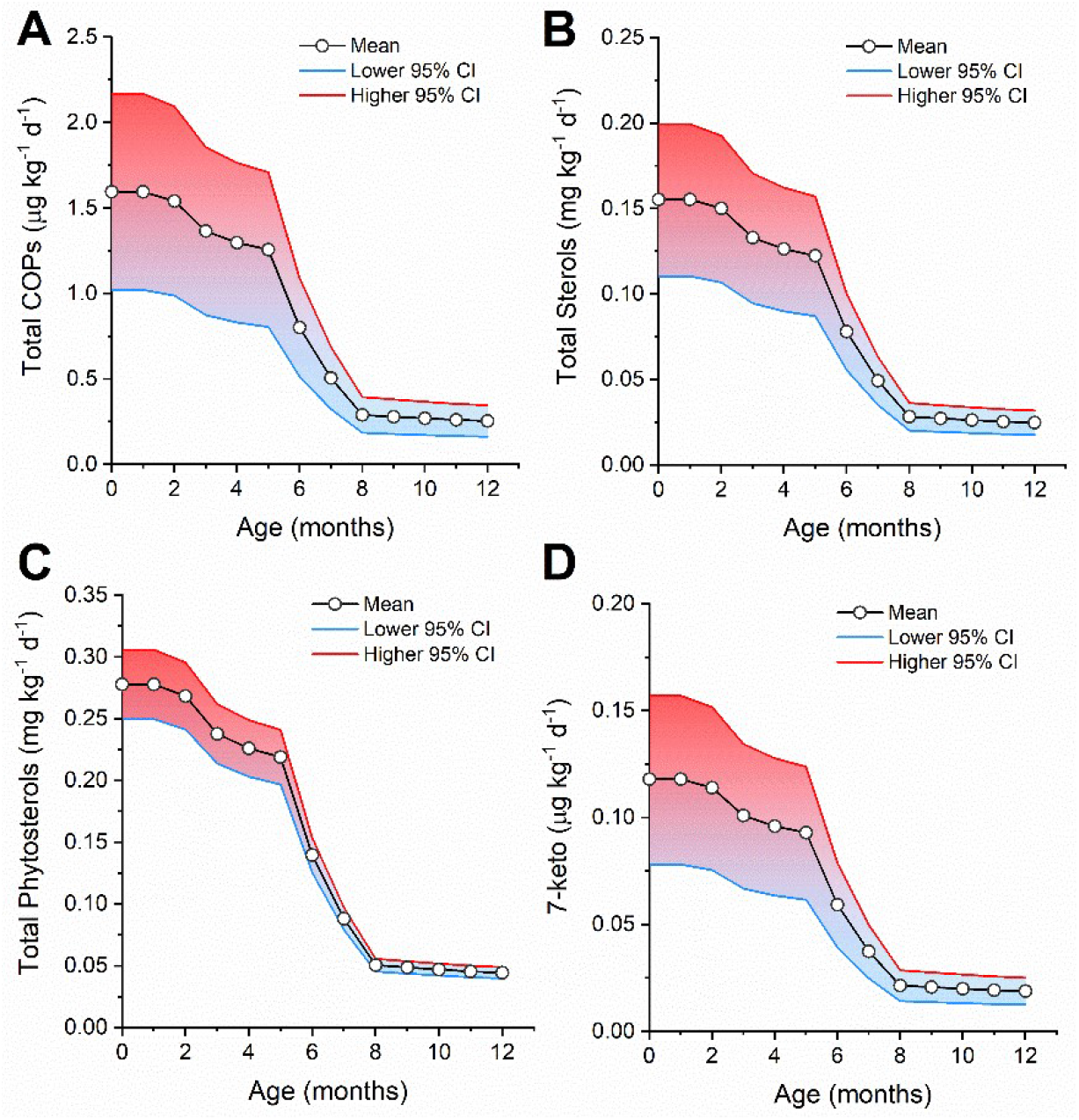
Dietary intake estimation for **A)** total COPs, **B)** total sterols, **C)** total phytosterols, and **D)** 7-keto during the first year of infant development.

## Discussion

Diet plays a critical role not only in infants’ growth, body composition, and development of the immune system, but also in the potential risk of developing metabolic disorders and obesity in adulthood (Lemaire, Le Huerou-Luron, & Blat, 2018). IFs provide nourishment at both macro- and micronutrient levels (Fuglestad et al., 2017), and although pediatric recommendations do not lean towards one formulation or another, as from the present study PIF and LIF have different macronutrient constituents. Powdered IF is put through manufacturing steps such as mixing, evaporation, and spray drying. PIF is usually stored in an aluminum can and vacuum sealed with nitrogen until the consumer first opens it (Jiang & Guo, 2014). Liquid IF can be either concentrated (needs to be mixed with water before consumption) or ready-to-fed and usually sterilization with high temperatures, followed by aseptic packaging are applied to guarantee microbial safety. In PIF, the high load in oxidized sterols and MDA hints a potential cause in the process of manufacturing. The LIFs undergo ultra-high temperature pasteurization at temperatures around 135-150ºC for less than 2.5 seconds, the IF is heated between 85°C and 120°C to emulsify the water and oil phases. Then the pH is adjusted, and some minor ingredients are added, while the PIF is spray dried using air at 170-250ºC with a product temperature >100ºC for longer times. Spray drying equipment can also provide indirect or direct heating, combined with low NOx (nitrogen oxides) or high NOx in the combustion gases which may affect the formation of oxidized derivatives (Chan et al., 1993). In addition, glass bottles, metal cans, polypropylene bottles, or cardboard with layers of polypropylene and aluminum foil are some materials commonly used for packaging of IF and their impact on the oxidative status is still under investigation (Guo, 2014).

It is well known that heat is one of the major triggers of lipid and cholesterol peroxidation (K. Al-Ismail, 2002; K. M. Al-Ismail, Herzallah, & Humied, 2007; Beltran, Pla, Capellas, Yuste, & Mor-Mur, 2004; Li et al., 1996; Mazalli & Bragagnolo, 2009; Medina-Meza, Rodriguez-Estrada, Lercker, Soto-Rodriguez, & Garcia, 2011; Smith, 1996). However, it is worth noting that the oxidative load is balanced by a higher antioxidant capacity due to the supplementation of antioxidants, particularly tocopherols. The balance between cholesterol and phytosterol intake is critical for infant development. Breast milk has 0.10-0.15 mg/mL of cholesterol, whereas cow-milk based infant formulas 0.01-0.04 mg/mL, meaning that infants alimented with milk formulation have less cholesterol intake (Wagner & von Stockhausen, 1988). Since the late ‘70s, it was known that milk formulas enriched with vegetable oils cause reduction of plasma cholesterol and LDL, with a concurrent increase of plasma and tissue phytosterols (M. Mellies, Glueck, C. J., Sweeney, C., Fallat, R. W., Tsang, R. C., Ishikawa, T. T., 1976; M. J. Mellies, Ishikawa, T. T., Glueck, C. J., Bove, K., Morrison, J., 1976). For instance, infants suffering sitosterolemia have increased absorption of phytosterols and cholesterol, causing accelerated atherosclerosis (Yoo, 2016). Higher intake of phytosterols is clearly associated with parenteral nutrition-associated liver disease (Kurvinen et al., 2012), which can be irreversible and devastating for infants (Teitelbaum & Tracy, 2001). A recent work on absorption kinetics, showed that neonates rapidly accumulate sitosterol and campesterol when supplied with soybean oil lipids (Nghiem-Rao et al., 2015). Thus, our large database could support further clinical studies and personalized nutrition for individuals possessing genetic deficiencies that lead to uncontrolled sterols accumulation.

LIFs contain higher amounts of COPs, although differences in individual COPs were not significant **(Table 1)** given the broad distribution across samples. However, 7-keto was significantly higher in PIF (p<0.01), thus we propose this compound as a potential biomarker of formula processing. 7-keto was already proposed as a biomarker of cholesterol oxidation in food matrices (Rodriguez-Estrada et al., 2014). Dehydrated products as milk powder seem to be particularly susceptible to generate 7-keto from cholesterol autoxidation (Sieber, 2005), which is possibly due to the high temperature that favors the conversion from both 5,6-epoxides epimers and 7-hydroxy precursors (Nourooz-Zadeh & Appelqvist, 1988). 7-keto is one of the most studied COPs, due to several biological effects reported either in cell or animal studies. Regarding infants, 7-keto is a biomarker for the early detection of inherited disorders related to cholesterol metabolism, including Niemann-Pick Type C disease and acid sphingomyelinase disease (Boenzi et al., 2016; Lin et al., 2014). Thus, it is important to know that formula intake can possibly result in an increase of 7-keto plasma concentration; however, we are far from understanding the rate and extent of COPs adsorption in infants.

Finally, our study allowed a re-classification of the IF based on the metabolite signature, rather than label information **(Figure 3)**. Three groups emerged, having dominant determinants of COPs and phytosterol contents. We are aware that this classification is not exhaustive since it takes into consideration only lipid and steroid molecules. However, it is a starting point for further studies targeted to expand our knowledge in food composition, towards a data-driven approach for personalized nutrition.

## Conclusions

IF are a useful alternative to breast milk; however, the effects of ultra-processing, such as high temperatures may impact the redox balance (antioxidant vs. oxidant load), significantly affecting their nutritional quality. The balance between cholesterol and phytosterols should be evaluated pre and post-processing) to avoid oxidized products and ensure optimal cholesterol absorption on infant’s diet, which is vital during the first months of life. In addition, the nutritional quality of IF may be compromised if the microbial reduction is the only standard for ensuring food safety. Lipids and proteins are thermally sensitive molecules which may generate hundreds of oxidized derivatives, harmful to human health. Enhancing the manufacturing processes, especially for PIF, should be implemented to reduce the generation of oxidized compounds. More effective and stable antioxidants should also be added to lower the potential for oxidation. The redox balance (e.g., ToAC vs TBARs) of the final product should be a critical parameter to be evaluated as a hallmark of the overall nutritional quality of IF. In addition, the study of oxidized derivatives interaction with the gut microbiome is needed, especially on infants where the gut is not fully developed.

## Data Availability

The data that support the findings of this study are available from the corresponding author, IGMM, upon reasonable request.

## Acknowledgements

This study has been funded by SHORA Spectrum Health through the grant RC108332 and the USDA National Institute of Food and Agriculture, Hatch project MICL02526 to Alice Kilvington, and Michigan State University start-up funding to I.G.M.M. Many thanks to Karen Ferguson RD, CSP, Pediatric Intensive Care Unit & Pediatric Burn Service, Helen DeVos Children’s Hospital for her support and guidance.

