## Supplemental Information for "Lipidomics and Dietary Assessment of Infant Formulas Reveal High Intakes of Major Cholesterol Oxidative Product (7-ketocholesterol)"

**Table S1**. Nutritional composition of Infant formulations (IFs) used in this study.

| **Type** | **IF ID** | **Formula Name** | **Protein (g)** | **Fat (g)** | **CHO (g)** | **Vitamin A** |
| --- | --- | --- | --- | --- | --- | --- |
| Powder | PIF1 | Enfamil A.R. | 2.5 | 5.1 | 11.3 | 300 |
|  | PIF2 | Enfamil Prosobee | 2.5 | 5.3 | 10.6 | 300 |
|  | PIF3 | Similac Advance | 2.07 | 5.6 | 10.5 | 300 |
|  | PIF4 | Similac Sensitive | 2.1 | 5.4 | 11.1 | 300 |
|  | PIF5 | Similac Neosure | 2.8 | 5.5 | 10.1 | 350 |
|  | PIF6 | Gerber Good Start Gentle | 2.2 | 5.1 | 11.2 | 300 |
|  | PIF7 | Little Journey Infant Formula | 2.1 | 5.3 | 11 | 300 |
|  | PIF8 | Enfacare | 2.8 | 5.3 | 10.4 | 450 |
|  | PIF9 | Enfamil Gentlease | 2.3 | 5.3 | 10.8 | 300 |
|  | PIF10 | Neocate Infant | 2.8 | 5.1 | 10.8 | 280 |
|  | PIF11 | Elecare Infant | 3.1 | 4.8 | 10.7 | 273 |
|  | PIF12 | Similac Alimentum | 2.75 | 5.54 | 10.2 | 300 |
|  | PIF13 | Similac Soy Isomil | 2.45 | 5.46 | 10.4 | 300 |
|  | PIF14 | Enfamil Premium | 2 | 5.3 | 11.3 | 300 |
|  | PIF15 | Nutramigen | 2.8 | 5.3 | 10.3 | 300 |
| Liquid | LIF1 | Enfamil Prosobee | 2.5 | 5.3 | 10.6 | 300 |
|  | LIF2 | Enfamil Infant (concentrate) | 2 | 5.3 | 11.3 | 300 |
|  | LIF3 | Similac Advance | 2.07 | 5.4 | 11 | 300 |
|  | LIF4 | Similac Neosure | 2.8 | 5.5 | 10.1 | 350 |
|  | LIF5 | Gerber Good Start Gentle | 2.2 | 5.1 | 11.6 | 300 |
|  | LIF6 | Pregestimil | 2.8 | 5.6 | 10.2 | 350 |
|  | LIF7 | Enfamil 24 | 2.1 | 5.3 | 11.2 | 300 |
|  | LIF8 | Enfamil Premature High Protein 24 Cal | 3.6 | 5 | 10.5 | 1350 |
|  | LIF9 | Enfamil Premature Formula, with Iron 20 Cal | 3.3 | 5 | 10.8 | 1350 |
|  | LIF10 | Enfamil Gentlease | 2.3 | 5.3 | 10.8 | 300 |
|  | LIF11 | Similac Alimentum | 2.75 | 5.54 | 10.2 | 300 |
|  | LIF12 | Enfamil Premium | 2 | 5.3 | 11.3 | 300 |
|  | LIF13 | Nutramigen | 2.8 | 5.3 | 10.3 | 300 |
|  | LIF14 | Enfaport | 3.5 | 5.5 | 10 | 350 |
|  | LIF15 | Enfacare | 2.8 | 5.3 | 10.4 | 450 |

In the label, fat, protein, and carbohydrates (CHO) values are reported in g per serving; vitamins as IU per 100 calories.

**Table S2**. Fat, protein, fatty acid, COPs, TBARS and ToAC content in IFs.

| **ID** | **Fat** | **Protein** | **Squalene** | **Desmosterol** | **SFA** | **MUFA** | **PUFA** | **TBARs** | **ToAC** |
| --- | --- | --- | --- | --- | --- | --- | --- | --- | --- |
| PIF1 | 1.42 ± 0.21^a^ | 12.1 ± 0.0^g,h^ | 0.092 ± 0.021^a,b,c,d^ | 0.204 ± 0.058^a,b^ | 41.28 ± 0.27^c,d,e,f^ | 34.32 ± 4.94^d,e,f,g^ | 24.39 ± 5.22 ^b,c,d,e,f,g,h,i^ | 0.043 ± 0.015 | 1.89 ± 0.47^a,b^ |
| PIF2 | 2.42 ± 0.52^a,b^ | 12.4 ± 0.1^h^ | 0.191 ± 0.020^d^ | 0.406 ± 0.039^b^ | 42.91 ± 1.38^e,f^ | 36.60 ± 0.86^f,g,h,i,j^ | 20.49 ± 0.52^b,c,d^ | 0.105 ± 0.06 | 3.94 ± 1.68^a,b^ |
| PIF3 | 3.11 ± 1.05^a,b^ | 10.9 ± 0.0^f,g^ | ND | 0.228 ± 0.126^a,b^ | 31.75 ± 0.76^a,b^ | 41.66 ± 0.47^j,k,l^ | 26.59 ± 0.28 ^h,i^ | 0.054 ± 0.002 | 2.85 ± 1.64^a,b^ |
| PIF4 | 2.43 ± 0.57^a,b^ | 11.0 ± 0.1^e,f^ | ND | 0.178 ± 0.033^a,b^ | 32.80 ± 0.37^a,b^ | 41.70 ± 0.33^j,k,l^ | 25.49 ± 0.04 ^d,e,f,g,h,i^ | 0.044 ± 0.000 | 2.00 ± 0.39^a,b^ |
| PIF5 | 2.57 ± 0.08^a,b^ | 14.7 ± 0.1^k^ | 0.013 ± 0.004^a^ | 0.175 ± 0.026^a,b^ | 37.59 ± 0.26^b,c,d,e^ | 36.61 ± 0.23^f,g,h,i,j^ | 25.79 ± 0.02 ^e,f,g,h,i^ | 0.087 ± 0.032 | 33.09 ± 1.85^d^ |
| PIF6 | 2.41 ± 0.65^a,b^ | 10.8 ± 0.1^e,f^ | 0.111 ± 0.003^a,b,c,d^ | 0.208 ± 0.123^a,b^ | 41.98 ± 1.27^d,e,f^ | 30.57 ± 0.39^c,d^ | 27.45 ± 0.87^i^ | 0.016 ± 0.013 | 5.26 ± 1.02^a,b,c^ |
| PIF7 | 2.69 ± 0.98 ^a,b^ | 11.0 ± 0.0^e,f^ | 0.049 ± 0.016^a,b,c^ | 0.338 ± 0.072^a,b^ | 42.70 ± 0.32^e,f^ | 36.03 ± 0.09^e,f,g,h,i^ | 21.27 ± 0.24^b,c,d,e,f,g^ | 0.067 ± 0.032 | 30.98 ± 3.74^d^ |
| PIF8 | 3.55 ± 0.86^b^ | 11.7 ± 0.1^f,g^ | 0.177 ± 0.046^c,d^ | 0.396 ± 0.109^b^ | 41.41 ± 0.12^c,d,e,f^ | 38.32 ± 0.07^g,h,i,j,k^ | 20.27 ± 0.05^b,c,d^ | 0.034 ± 0.005 | 8.40 ± 1.22^a,b,c^ |
| PIF9 | 3.53 ± 0.57^b^ | 14.7 ± 0.1^k,l^ | 0.024 ± 0.009^a,b^ | 0.402 ± 0.132^b^ | 36.05 ± 1.15^b,c,d^ | 42.49 ± 0.64^k,l^ | 21.46 ± 0.51^b,c,d,e,f,g,h^ | 0.076 ± 0.053 | 7.79 ± 1.02^a,b,c^ |
| PIF10 | 2.48 ± 0.30 ^a,b^ | 15.3 ± 0.1^l^ | 0.006 ± 0.002^a^ | 0.402 ± 0.132^a,b^ | 36.10 ± 1.01^b,c,d^ | 38.71 ± 1.24^g,h,i,j,k^ | 25.19 ± 0.22 ^c,d,e,f,g,h,i^ | 0.059 ± 0.004 | 6.45 ± 1.74^a,b,c^ |
| PIF11 | 2.98 ± 0.28 ^a,b^ | 14.2 ± 0.1^j,k^ | 0.028 ± 0.023^a,b^ | 0.182 ± 0.007^a,b^ | 40.83 ± 0.43^c,d,e,f^ | 34.87 ± 0.27^d,e,f,g,h^ | 24.60 ± 0.16 ^b,c,d,e,f,g,h,i^ | 0.040 ± 0.094 | 8.77 ± 1.04^a,b,c^ |
| PIF12 | 2.81 ± 0.16 ^a,b^ | 13.2 ± 0.1^i^ | 0.006 ± 0.002^a^ | 0.138 ± 0.022^a,b^ | 29.27 ± 0.19^a^ | 44.65 ± 0.60^k^ | 26.08 ± 0.41 ^f,g,h,i^ | 0.082 ± 0.020 | 9.41 ± 3.69^b,c^ |
| PIF13 | 2.36 ± 0.83 ^a,b^ | 10.4 ± 0.1^e^ | 0.088 ± 0.037^a,b,c,d^ | 0.317 ± 0.121^a,b^ | 41.33 ± 0.02^c,d,e,f^ | 37.90 ± 0.05^g,h,i,j,k^ | 20.77 ± 0.03^b,c,d,e^ | 0.043 ± 0.028 | 8.61 ± 7.37^a,b,c^ |
| PIF14 | 2.75 ± 0.14 ^a,b^ | 13.7 ± 0.1^i,j^ | 0.108 ± 0.006^a,b,c,d^ | 0.305 ± 0.005^a,b^ | 39.37 ± 0.33^c,d,e,f^ | 40.83 ± 0.32^i,j,k,l^ | 19.79 ± 0.00^b^ | 0.035 ± 0.017 | 25.10 ± 2.14^d^ |
| PIF15 | 2.66 ± 0.65 ^a,b^ | 13.9 ± 0.0^j^ | 0.012 ± 0.006^a^ | 0.272 ± 0.034^a,b^ | 39.55 ± 3.60^c,d,e,f^ | 39.67 ± 2.69^h,i,j,k,l^ | 20.77 ± 0.91^b,c,d,e^ | 0.122 ± 0.059 | 5.51 ± 0.91^a,b,c^ |
| LIF1 | 1.95 ± 0.13 ^a,b^ | 1.3 ± 0.3^a,b^ | 0.100 ± 0.022^a,b,c,d^ | 0.232 ± 0.010^a,b^ | 42.30 ± 0.12^d,e,f^ | 37.03 ± 0.17^g,h,i,j^ | 20.67 ± 0.05^b,c,d,e^ | 0.036 ± 0.009 | 0.80 ± 0.35^a^ |
| LIF2 | 2.3 ± 0.62 ^a,b^ | 2.9 ± 0.4^d^ | 0.101 ± 0.018^a,b,c,d^ | 0.246 ± 0.010^a,b^ | 42.99 ± 0.02^e,f^ | 36.57 ± 0.12^f,g,h,i,j^ | 20.43 ± 0.14^b,c,d^ | 0.072 ± 0.037 | 3.34 ± 1.37^a,b^ |
| LIF3 | 2.30 ± 0.62 ^a,b^ | 1.1 ± 0.3^a^ | ND | 0.198 ± 0.013^a,b^ | 32.49 ± 0.58^a,b^ | 41.08 ± 0.73^i,j,k,l^ | 26.42 ± 1.31 ^g,h,i^ | 0.068 ± 0.037 | 0.96 ± 0.21^a^ |
| LIF4 | 2.40 ± 0.06 ^a,b^ | 2.0 ± 0.3^c^ | 0.088 ± 0.059^a,b,c,d^ | 0.087 ± 0.025^a^ | 63.56 ± 0.05^h^ | 0.91 ± 0.01^a^ | 35.53 ± 0.05^j^ | 0.042 ± 0.038 | 0.86 ± 0.02^a^ |
| LIF5 | 2.34 ± 0.20 ^a,b^ | 1.5 ± 0.0^a,b,c^ | 0.092 ± 0.028^a,b,c,d^ | 0.151 ± 0.015^a,b^ | 45.70 ± 6.33^f^ | 28.55 ± 3.31^c^ | 25.75 ± 3.02 ^e,f,g,h,i^ | 0.023 ± 0.004 | 5.38 ± 0.73^a,b,c^ |
| LIF6 | 2.39 ± 0.07 ^a,b^ | 2.1 ± 0.3^c^ | ND | 0.258 ± 0.058^a,b^ | 54.99 ± 2.14^g^ | 16.80 ± 0.83^b^ | 28.20 ± 1.31^i^ | 0.018 ± 0.007 | 8.96 ± 2.01^a,b,c^ |
| LIF7 | 2.56 ± 0.28 ^a,b^ | 1.5 ± 0.3^a,b,c^ | 0.151 ± 0.068^b,c,d^ | 0.224 ± 0.112^a,b^ | 43.00 ± 0.10^e,f^ | 36.76 ± 0.07^f,g,h,i,j^ | 20.24 ± 0.03^b,c^ | 0.043 ± 0.007 | 1.21 ± 0.19^a,b^ |
| LIF8 | 1.95 ± 0.16 ^a,b^ | 2.8 ± 0.2^d^ | ND | 0.221 ± 0.097^a,b^ | 44.66 ± 0.09^f^ | 31.75 ± 0.11^c,d,e,f^ | 23.59 ± 0.02 ^b,c,d,e,f,g,h,i^ | 0.040 ± 0.015 | 2.38 ± 1.43^a,b^ |
| LIF9 | 1.9 ± 0.10 ^a,b^ | 1.9 ± 0.4^a,b^ | ND | 0.229 ± 0.083^a,b^ | 44.61 ± 0.97^f^ | 31.11 ± 0.47^c,d,e^ | 24.28 ± 0.50 ^b,c,d,e,f,g,h,i^ | 0.077 ± 0.021 | 1.39 ± 0.44^a,b^ |
| LIF10 | 2.72 ± 0.40 ^a,b^ | 1.6 ± 0.0^a,b,c^ | 0.102 ± 0.008^a,b,c,d^ | 0.228 ± 0.022^a,b^ | 42.61 ± 0.39^e,f^ | 37.66 ± 0.22^g,h,i,j,k^ | 19.72 ± 0.17^b^ | 0.056 ± 0.001 | 7.87 ± 1.18^a,b,c^ |
| LIF11 | 1.96 ± 0.17 ^a,b^ | 1.9 ± 0.0^b,c^ | ND | 0.131 ± 0.015^a,b^ | 35.09 ± 2.64^a,b,c^ | 12.31 ± 0.47^b^ | 52.60 ± 2.17^k^ | 0.019 ± 0.007 | 12.44 ± 1.12^c^ |
| LIF12 | 2.48 ± 0.73 ^a,b^ | 1.3 ± 0.0^a,b^ | 0.172 ± 0.081^c,d^ | 0.243 ± 0.109^a,b^ | 41.69 ± 0.53^d,e,f^ | 37.27 ± 0.28^g,h,i,j,k^ | 21.04 ± 0.82^b,c,d,e,f^ | 0.047 ± 0.022 | 1.08 ± 0.17^a^ |
| LIF13 | 2.89 ± 0.30 ^a,b^ | 1.2 ± 0.0^a^ | 0.060 ± 0.085^a,b,c^ | 0.307 ± 0.032^a,b^ | 64.65 ± 0.67^h^ | 0.42 ± 0.01^a^ | 34.93 ± 0.68^j^ | 0.125 ± 0.048 | 12.73 ± 1.37^c^ |
| LIF14 | 3.03 ± 0.30 ^a,b^ | 3.4 ± 0.1^d^ | ND | 0.237 ± 0.103^a,b^ | 85.06 ± 0.10^i^ | 4.26 ± 0.06^a^ | 10.68 ± 0.05^a^ | 0.046 ± 0.005 | 3.19 ± 1.37^a,b^ |
| LIF15 | 3.30 ± 0.68 ^a,b^ | 2.1 ± 0.0^c^ | ND | 0.185 ± 0.015^a,b^ | 40.09 ± 0.19^c,d,e,f^ | 36.92 ± 0.08^f,g,h,i,j^ | 22.99 ± 0.11^b,c,d,e,f,g,h,i^ | 0.099 ± 0.042 | 1.27 ± 0.19^a,b^ |

Squalene, desmosterol and total sterols values reported as mg per scoop (9 grams) of powder or per liquid equivalent to1 ready to feed bottle (60 mL). Fat, protein, SFA, MUFA and PUFA are reported in %. TBARs values reported as ug and ToAC reported as mg/L. Different subscripts letters indicate significant difference (p < 0.05).

**Table S3.** Sterols, phytosterols and squalene content in IFs.

| **Type** | **Sample** | **Cholesterol** | **Desmosterol** | **Total Sterols** | **Stigmasterol** | **β-Sitosterol** | **Campesterol** | | **Fucosterol** | | **Total phytosterols** | |
| --- | --- | --- | --- | --- | --- | --- | --- | --- | --- | --- | --- | --- |
| Powder | PIF1 | 0.443 ± 0.132_a,b,c,d_ | 0.204 ± 0.058^a,b^ | 0.647±0.075 | 0.185 ± 0.017^a^ | 1.076 ± 0.029^a,b^ | | 0.287 ± 0.059^a^ | | 0.076 ± 0.006^a,b^ | | 1.624±0.098 |
|  | PIF2 | 0.089 ± 0.125^a.b^ | 0.406 ± 0.039^b^ | 0.495±0.164 | 0.387 ± 0.012^a,b^ | 2.478 ± 0.395^b,c,d^ | | 0.603 ± 0.037^a,b,c^ | | 0.197 ± 0.070^a,b,c^ | | 2.853±0.490 |
|  | PIF3 | 2.220 ± 0.758^a,b,c,d,e,f,g^ | 0.228 ± 0.126^a,b^ | 2.448±0.884 | 0.569 ± 0.250^b^ | 3.801 ± 1.365^b,c,d^ | | 0.909 ± 0.408^b,c^ | | 0.359 ± 0.195^c^ | | 3.666±2.217 |
|  | PIF4 | 0.513 ± 0.521^a,b,c,d,e^ | 0.178 ± 0.033^a,b^ | 0.690±0.554 | 0.393 ± 0.091_a,b_ | 2.460 ± 0.717^d^ | | 0.642 ± 0.159^a,b,c^ | | 0.253 ± 0.115^b,c^ | | 5.262±1.082 |
|  | PIF5 | 2.481 ± 0.455^a,b,c,d,e,f,g^ | 0.175 ± 0.026^a,b^ | 2.656±0.430 | 0.413 ± 0.022^a,b^ | 2.244 ± 0.064^b,c,d^ | | 0.565 ± 0.018^a,b^ | | 0.241 ± 0.015^b,c^ | | 5.637±0.009 |
|  | PIF6 | 3.325 ± 0.158^f,g^ | 0.208 ± 0.123^a,b^ | 3.532±0.281 | 0.584 ± 0.284^b^ | 2.33 ± 0.847^b,c,d^ | | 0.755 ± 0.346^a,b,c^ | | 0.183 ± 0.059^a,b,c^ | | 4.291±1.537 |
|  | PIF7 | 1.974 ± 0.689^a,b,c,d,e,f,g^ | 0.338 ± 0.072^a,b^ | 2.312±0.762 | 0.359 ± 0.100^a,b^ | 2.015 ± 0.453^a,b,c,d^ | | 0.576 ± 0.230^a,b^ | | 0.169 ± 0.060^a,b,c^ | | 3.747±0.844 |
|  | PIF8 | 3.222 ± 0.640^e,f,g^ | 0.396 ± 0.109^b^ | 4.084±0.953 | 0.340 ± 0.100^a,b^ | 2.011 ± 0.636^a,b,c,d^ | | 0.511 ± 0.146^a,b^ | | 0.104 ± 0.040^a,b^ | | 3.226±0.852 |
|  | PIF9 | 0.089 ± 0.018^a,b^ | 0.402 ± 0.132^b^ | 3.618±0.749 | 0.241 ± 0.043^a,b^ | 3.313 ± 0.708^c,d^ | | 1.170 ± 0.273^c^ | | 0.212 ± 0.042^a,b,c^ | | 3.463±0.922 |
|  | PIF10 | 0.151 ± 0.078^a,b,c^ | 0.402 ± 0.132^a,b^ | 0.491±0.150 | 0.247 ± 0.015^a,b^ | 1.432 ± 0.285^a,b^ | | 0.431 ± 0.118^a,b^ | | 0.162 ± 0.042^a,b,c^ | | 4.199±1.066 |
|  | PIF11 | 0.041 ± 0.058^a^ | 0.182 ± 0.007^a,b^ | 0.333±0.021 | 0.439 ± 0.043^a,b^ | 2.175 ± 0.169^a,b,c,d^ | | 0.671 ± 0.050^a,b,c^ | | 0.215 ± 0.004^a,b,c^ | | 3.854±0.431 |
|  | PIF12 | 0.099 ± 0.072^a,b,c^ | 0.138 ± 0.022^a,b^ | 0.222±0.051 | 0.420 ± 0.096^a,b^ | 2.079 ± 0.147^a,b,c,d^ | | 0.553 ± 0.095^a,b^ | | 0.229 ± 0.023^a,b,c^ | | 2.645±0.258 |
|  | PIF13 | 1.477 ± 0.424^a,b,c,d,e,f^ | 0.317 ± 0.121^a,b^ | 0.236±0.094 | 0.358 ± 0.140^a,b^ | 1.792 ± 0.610^a,b,c^ | | 0.498 ± 0.220^a,b^ | | 0.094 ± 0.018^a,b^ | | 3.120±0.315 |
|  | PIF14 | 0.165 ± 0.021^a,b,c^ | 0.305 ± 0.005^a,b^ | 1.794±0.303 | 0.280 ± 0.037^a,b^ | 1.455 ± 0.118^a,b^ | | 0.389 ± 0.039^a,b^ | | 0.072 ± 0.035^a,b^ | | 3.642±0.988 |
|  | PIF15 | 3.812 ± 0.920^f,g^ | 0.272 ± 0.034^a,b^ | 0.469±0.026 | 0.397 ± 0.096^a,b^ | 2.010 ± 0.633^a,b,c,d^ | | 0.442 ± 0.098^a,b^ | | 0.115 ± 0.025^a,b^ | | 2.965±0.229 |
| Liquid | LIF1 | 0.014 ± 0.020^a^ | 0.232 ± 0.010^a,b^ | 0.246±0.011 | 0.256 ± 0.026^a,b^ | 1.417 ± 0.083^a,b^ | | 0.304 ± 0.005^a^ | | 0.103 ± 0.047^a,b^ | | 2.990±0.067 |
|  | LIF2 | 1.540 ± 0.234^a,b,c,d,e,f^ | 0.246 ± 0.010^a,b^ | 1.785±0.124 | 0.262 ± 0.021^a,b^ | 1.198 ± 0.124^a,b^ | | 0.340 ± 0.001^a,b^ | | 0.115 ± 0.031^a,b^ | | 2.966±1.004 |
|  | LIF3 | 1.295 ± 0.072^a,b,c,d,e,f^ | 0.198 ± 0.013^a,b^ | 1.493±0.059 | 0.340 ± 0.021^a,b^ | 2.097 ± 0.220^a,b,c,d^ | | 0.604 ± 0.012^a,b,c^ | | 0.141 ± 0.035^a,b,c^ | | 4.002±0.288 |
|  | LIF4 | 1.696 ± 0.546^a,b,c,d,e,f^ | 0.087 ± 0.025^a^ | 1.782±0.571 | 0.273 ± 0.068^a,b^ | 0.988 ± 0.208^a,b^ | | 0.353 ± 0.058^a,b^ | | 0.049 ± 0.004^a,b^ | | 4.936±0.329 |
|  | LIF5 | 2.913 ± 0.185^d,e,f,g^ | 0.151 ± 0.015^a,b^ | 3.063±0.170 | 0.271 ± 0.032^a,b^ | 1.164 ± 0.923^a,b^ | | 0.359 ± 0.041^a,b^ | | 0.137 ± 0.042^a,b,c^ | | 3.379±0.201 |
|  | LIF6 | 0.012 ± 0.017^a^ | 0.258 ± 0.058^a,b^ | 0.269±0.041 | 0.272 ± 0.048^a,b^ | 0.923 ± 0.115^a,b^ | | 0.316 ± 0.057^a^ | | 0.050 ± 0.011^a,b^ | | 2.272±0.231 |
|  | LIF7 | 2.487 ± 0.413^a,b,c,d,e,f,g^ | 0.224 ± 0.112^a,b^ | 2.711±0.525 | 0.301 ± 0.017^a,b^ | 1.610 ± 0.035^a,b,c^ | | 0.093 ± 0.625^a^ | | 0.183 ± 0.104^a,b,c^ | | 2.642±0.041 |
|  | LIF8 | 3.241 ± 0.104^e,f,g^ | 0.221 ± 0.097^a,b^ | 3.462±0.201 | 0.269 ± 0.074^a,b^ | 1.284 ± 0.169^a,b^ | | 0.265 ± 0.105^a^ | | 0.175 ± 0.113^a,b,c^ | | 3.499±0.251 |
|  | LIF9 | 2.831 ± 0.039^c,d,e,f,g^ | 0.229 ± 0.083^a,b^ | 3.060±0.122 | 0.162 ± 0.019^a^ | 0.918 ± 0.083^a,b^ | | 0.234 ± 0.045^a^ | | 0.058 ± 0.001^a,b^ | | 3.593±0.147 |
|  | LIF10 | 3.817 ± 1.293^f,g^ | 0.228 ± 0.022^a,b^ | 4.045±1.315 | 0.280 ± 0.028^a,b^ | 1.247 ± 0.135^a,b^ | | 0.277 ± 0.010^a^ | | 0.076 ± 0.001^a,b^ | | 3.280±0.173 |
|  | LIF11 | 0.066 ± 0.032^a,b^ | 0.131 ± 0.015^a,b^ | 0.197±0.046 | 0.249 ± 0.037^a,b^ | 1.139 ± 0.191^a,b^ | | 0.270 ± 0.075^a^ | | 0.112 ± 0.006^a,b^ | | 2.550±0.309 |
|  | LIF12 | 0.124 ± 0.112^a,b,c^ | 0.243 ± 0.109^a,b^ | 0.367±0.221 | 0.279 ± 0.106^a,b^ | 1.342 ± 0.505^a,b^ | | 0.333 ± 0.145^a,b^ | | 0.138 ± 0.041^a,b,c^ | | 2.742±0.798 |
|  | LIF13 | 1.796 ± 1.396^a,b,c,d,e,f,g^ | 0.307 ± 0.032^a,b^ | 1.719±1.428 | 0.348 ± 0.033^a,b^ | 1.753 ± 0.215^a,b,c^ | | 0.461 ± 0.063^a,b^ | | 0.171 ± 0.009^a,b,c^ | | 2.737±0.302 |
|  | LIF14 | 2.802 ± 1.052^b,c,d,e,f,g^ | 0.237 ± 0.103^a,b^ | 3.039±1.155 | 0.109 ± 0.017^a^ | 0.380 ± 0.137^a^ | | 0.166 ± 0.034^a^ | | 0.092 ± 0.011^a,b^ | | 2.196±0.177 |
|  | LIF15 | 4.492 ± 2.205^g^ | 0.185 ± 0.015^a,b^ | 4.676±2.190 | 0.252 ± 0.030^a,b^ | 1.316 ± 0.055^a,b^ | | 0.303 ± 0.035^a^ | | ND | | 2.358±0.061 |

Values reported as mg per scoop (9 grams of powder) or per liquid equivalent to 1 ready to feed bottle (60 mL). Different letters within columns indicate significant differences (p < 0.05).

**Table S4.** Tocopherols profile of liquid and powder IF.

| **Type** | **Sample** | **α-Tocopherol** | **β-Tocopherol** | **γ-Tocopherol** | **δ-Tocopherol** | **Total Tocopherols** |
| --- | --- | --- | --- | --- | --- | --- |
| Powder | PIF1 | 0.081 ± 0.028 | ND | 0.029 ± 0.019 | 0.062 ± 0.007^a^ | 0.172 ± 0.040^a,b^ |
|  | PIF2 | 0.014 ± 0.017 | ND | 0.020 ± 0.001 | 0.120 ± 0.050^a^ | 0.155 ± 0.077^a,b^ |
|  | PIF3 | ND | 0.018 ± 0.008^a,b^ | 0.064 ± 0.037 | 0.451 ± 0.203^a,b^ | 0.532 ± 0.248^b,c^ |
|  | PIF4 | 0.016 ± 0.016 | 0.004 ± 0.001^a,b^ | 0.029 ± 0.023 | 0.276 ± 0.104^a,b^ | 0.325 ± 0.144^a,b,c^ |
|  | PIF5 | 0.055 ± 0.039 | 0.029 ± 0.030^a,b^ | 0.062 ± 0.050 | 0.278 ± 0.030^a,b^ | 0.424 ± 0.049^a,b,c^ |
|  | PIF6 | 0.030 ± 0.025 | 0.044 ± 0.051^a,b^ | 0.056 ± 0.068 | 0.613 ± 0.252^b^ | 0.743 ± 0.397^c^ |
|  | PIF7 | 0.017 ± 0.006 | 0.020 ± 0.005^a,b^ | 0.028 ± 0.017 | 0.171 ± 0.045^a,b^ | 0.237 ± 0.060^a,b^ |
|  | PIF8 | 0.036 ± 0.043 | 0.010 ± 0.010^a,b^ | 0.022 ± 0.021 | 0.098 ± 0.035^a^ | 0.166 ± 0.109^a,b^ |
|  | PIF9 | 0.015 ± 0.012 | 0.012 ± 0.010^a,b^ | 0.017 ± 0.006 | 0.027 ± 0.015^a^ | 0.071 ± 0.030^a,b^ |
|  | PIF10 | 0.025 ± 0.003 | 0.010 ± 0.003^a,b^ | 0.009 ± 0.001 | 0.110 ± 0.000^a^ | 0.155 ± 0.005^a,b^ |
|  | PIF11 | 0.037 ± 0.006 | 0.015 ± 0.006^a,b^ | 0.034 ± 0.004 | 0.294 ± 0.012^a,b^ | 0.380 ± 0.016^a,b,c^ |
|  | PIF12 | 0.021 ± 0.016 | 0.023 ± 0.009^a,b^ | 0.059 ± 0.020 | 0.262 ± 0.039^a,b^ | 0.364 ± 0.034^a,b,c^ |
|  | PIF13 | 0.026 ± 0.033 | 0.006 ± 0.005^a,b^ | 0.021 ± 0.018 | 0.101 ± 0.042^a^ | 0.154 ± 0.098^a,b^ |
|  | PIF14 | 0.046 ± 0.013 | 0.010 ± 0.001^a,b^ | 0.021 ± 0.007 | 0.116 ± 0.033^a^ | 0.193 ± 0.041^a,b^ |
|  | PIF15 | 0.058 ± 0.054 | 0.012 ± 0.005^a,b^ | 0.017 ± 0.002 | 0.082 ± 0.008^a^ | 0.169 ± 0.043^a,b^ |
| Liquid | LIF1 | 0.017 ± 0.006 | 0.027 ± 0.027^a,b^ | 0.014 ± 0.016 | 0.047 ± 0.022^a^ | 0.106 ± 0.028^a,b^ |
|  | LIF2 | 0.042 ± 0.048 | 0.006 ± 0.001^a,b^ | 0.000 ± 0.000 | 0.019 ± 0.003^a^ | 0.067 ± 0.050^a^ |
|  | LIF3 | 0.020 ± 0.003 | 0.004 ± 0.006^a,b^ | 0.000 ± 0.000 | 0.030 ± 0.042^a^ | 0.054 ± 0.051^a^ |
|  | LIF4 | 0.054 ± 0.076 | 0.097 ± 0.016^a,b^ | 0.000 ± 0.000 | 0.177 ± 0.003^a,b^ | 0.328 ± 0.096^a,b,c^ |
|  | LIF5 | 0.049 ± 0.013 | 0.048 ± 0.025^a,b^ | 0.051 ± 0.038 | 0.188 ± 0.011^a,b^ | 0.335 ± 0.036^a,b,c^ |
|  | LIF6 | 0.092 ± 0.043 | 0.055 ± 0.022^a,b^ | ND | 0.123 ± 0.045^a^ | 0.270 ± 0.110^a,b^ |
|  | LIF7 | 0.108 ± 0.081 | 0.040 ± 0.029^a,b^ | ND | 0.083 ± 0.062^a^ | 0.231 ± 0.172^a,b^ |
|  | LIF8 | 0.116 ± 0.020 | 0.035 ± 0.030^a,b^ | ND | 0.085 ± 0.023^a^ | 0.235 ± 0.034^a,b^ |
|  | LIF9 | 0.011 ± 0.011 | 0.000 ± 0.000^a^ | ND | 0.047 ± 0.002^a^ | 0.058 ± 0.013^a^ |
|  | LIF10 | 0.037 ± 0.009 | 0.006 ± 0.004^a,b^ | ND | 0.036 ± 0.004^a^ | 0.079 ± 0.017^a,b^ |
|  | LIF11 | 0.008 ± 0.003 | 0.006 ± 0.002^a,b^ | ND | 0.055 ± 0.004^a^ | 0.069 ± 0.009^a,b^ |
|  | LIF12 | 0.009 ± 0.005 | 0.009 ± 0.000^a,b^ | ND | 0.041 ± 0.011^a^ | 0.058 ± 0.015^a^ |
|  | LIF13 | 0.036 ± 0.022 | 0.099 ± 0.094^b^ | ND | 0.087 ± 0.069^a^ | 0.222 ± 0.142^a,b^ |
|  | LIF14 | 0.089 ± 0.021 | ND | ND | 0.144 ± 0.102^a^ | 0.233 ± 0.123^a,b^ |
|  | LIF15 | 0.094 ± 0.056 | ND | ND | 0.049 ± 0.000^a^ | 0.143 ± 0.056^a,b^ |

Values reported in mg per scoop (9 grams) of powder or per liquid equivalent to1 ready to feed bottle, (60 mL). Identical letters within columns indicate that samples are not statistically different, according to analysis of variance and Tukey’s mean comparison test (p < 0.05).

**Table S5.** Cholesterol Oxidation Products profile of liquid and powder IF.

| **Type** | **Sample** | **7α-OH** | **7β-OH** | **7-keto** | **25-OH** | **Triol** | **6-keto** | **5,6β-epoxy** | **5,6α-epoxy** | **Total COPs** |
| --- | --- | --- | --- | --- | --- | --- | --- | --- | --- | --- |
| Powder | PIF1 | 5.83 ± 2.37^a,b,c^ | 6.46 ± 3.57^a,b,c,d^ | 1.96 ± 0.68^a^ | 1.00 ± 0.31 | 0.86 ± 0.09 | 0.41 ± 0.09 | 0.30 ± 0.06^a,b^ | 0.06 ± 0.03^a^ | 16.95±6.31^a^ |
|  | PIF2 | 0.51 ± 0.18^a^ | 0.47 ± 0.17^a^ | 0.14 ± 0.03^a^ | 0.68 ± 0.01 | 0.48 ± 0.01 | 0.14 ± 0.09 | ND | ND | 2.41±0.50^a^ |
|  | PIF3 | 2.39 ± 1.75^a,b,c^ | 2.16 ± 1.88^a,b,c^ | 0.67 ± 0.57^a^ | 2.75 ± 1.06 | 1.03 ± 0.12 | 0.15 ± 0.11 | 0.24 ± 0.34^a,b^ | 0.11 ± 0.16^a^ | 10.52±1.44^ab^ |
|  | PIF4 | 2.89 ± 0.63^a,b,c^ | 3.05 ± 0.27^a,b,c^ | 1.78 ± 0.23^a^ | 2.33 ± 0.10 | 1.78 ± 0.17 | 0.68 ± 0.56 | 0.38 ± 0.05^a,b^ | 0.47 ± 0.20^a,b^ | 14.70±0.36^b^ |
|  | PIF5 | 5.86 ± 4.91^a,b,c^ | 6.08 ± 4.23^a,b,c,d^ | 2.36 ± 0.01^a,b^ | 2.10 ± 0.57 | 2.33 ± 0.11 | 0.73 ± 0.48 | 0.42 ± 0.09^a,b^ | 0.04 ± 0.06^a,b^ | 21.53±8.02^a^ |
|  | PIF6 | 4.94 ± 1.55^a,b,c^ | 6.76 ± 3.56^a,b,c,d^ | 5.50 ± 6.66^a,b^ | ND | 0.26 ± 0.36 | 14.44 ± 20.27 | 0.06 ± 0.08^a^ | 0.04 ± 0.06^a^ | 32.00±13.53^ab^ |
|  | PIF7 | 11.24 ± 5.34^a,b,c^ | 17.43 ± 2.26^b,c,d^ | 3.80 ± 4.22^a,b^ | 2.85 ± 1.20 | 2.29 ± 0.65 | 1.69 ± 1.80 | 0.32 ± 0.24^a,b^ | 0.38 ± 0.07^a,b^ | 40.83±0.28^a^ |
|  | PIF8 | 6.49 ± 0.22^a,b,c^ | 11.25 ± 4.34^a,b,c,d^ | 2.40 ± 1.33^a,b^ | 0.66 ± 0.93 | 0.65 ± 0.91 | 0.29 ± 0.19 | ND | ND | 21.73±4.24^a^ |
|  | PIF9 | 5.47 ± 0.33^a,b,c^ | 8.32 ± 0.50^a,b,c,d^ | 1.25 ± 0.00^a^ | 0.30 ± 0.42 | 0.43 ± 0.54 | 0.49 ± 0.14 | 0.08 ± 0.11^a^ | 0.07 ± 0.11^a^ | 16.42±0.86^a^ |
|  | PIF10 | 0.65 ± 0.06^a^ | 0.50 ± 0.03^a^ | 0.35 ± 0.03^a^ | 0.47 ± 0.20 | 0.52 ± 0.26 | 0.21 ± 0.14 | 0.03 ± 0.04^a^ | 0.07 ± 0.09^a^ | 2.80±0.67^a^ |
|  | PIF11 | 5.32 ± 1.76^a,b,c^ | 3.53 ± 1.24^a,b,c^ | 3.30 ± 1.74^a,b^ | ND | 0.83 ± 0.03 | 8.04 ± 10.46 | 0.81 ± 0.39^b,c^ | 1.39 ± 0.39^b^ | 25.79±19.33^ab^ |
|  | PIF12 | 1.43 ± 0.01^a,b^ | 2.17 ± 0.75^a,b,c^ | 1.23 ± 0.83^a^ | 0.45 ± 0.83 | 0.52 ± 0.74 | 1.04 ± 1.08 | 0.15 ± 0.21^a,b^ | 0.10 ± 0.15^a^ | 7.10±0.95^a^ |
|  | PIF13 | 2.96 ± 1.08^a,b,c^ | 1.96 ± 0.04^a,b^ | 2.00 ± 1.56^a^ | 2.08 ± 0.10 | 4.24 ± 5.62 | 0.40 ± 0.43 | 0.24 ± 0.20_­­_­^a,b^ | 0.24 ± 0.18^a^ | 15.18±8.89^ab^ |
|  | PIF14 | 5.44 ± 1.95^a,b,c^ | 6.57 ± 1.45^a,b,c,d^ | 1.60 ± 0.06^a^ | 0.56 ± 0.07 | 1.72 ± 0.96 | 0.36 ± 0.29 | 0.11 ± 0.06^a,b^ | 0.17 ± 0.05^a^ | 17.18±2.87^a^ |
|  | PIF15 | 1.96 ± 1.67^a,b,c^ | 2.02 ± 2.14^a,b^ | 1.26 ± 1.06^a^ | ND | 1.36 ± 0.25 | 0.15 ± 0.12 | 0.11 ± 0.02^a,b^ | 0.08 ± 0.04^a^ | 6.94±5.06^a^ |
| Liquid | LIF1 | 0.84 ± 0.63^a^ | 2.01 ± 1.68 ^a,b^ | 0.20 ± 0.28^a^ | ND | 0.70 ± 0.67 | ND | ND | ND | 3.75±1.26^a^ |
|  | LIF2 | 6.15 ± 3.48^a,b,c^ | 6.79 ± 3.74^a,b,c,d^ | 0.90 ± 0.27^a^ | ND | 0.57 ± 0.70 | 0.06 ± 0.08 | 0.08 ± 0.11^a^ | 0.03 ± 0.04^a^ | 14.58±8.43^a^ |
|  | LIF3 | 1.76 ± 0.72^a,b^ | 1.72 ± 0.41^a,b^ | 0.28 ± 0.00^a^ | ND | 0.53 ± 0.24 | 0.05 ± 0.00 | 0.06 ± 0.02^a^ | 0.06 ± 0.05^a^ | 4.46±2.46^a^ |
|  | LIF4 | 2.15 ± 0.40^a,b,c^ | 2.11 ± 0.37^a,b,c^ | 0.39 ± 0.24^a^ | ND | 0.57 ± 0.51 | ND | 0.15 ± 0.13^a,b^ | 0.13 ± 0.10^a^ | 5.51±0.96^a^ |
|  | LIF5 | 4.78 ± 1.51^a,b,c^ | 5.14 ± 1.09^a,b,c,d^ | 0.65 ± 0.01^a^ | 0.45 ± 0.19 | 0.60 ± 0.07 | 0.22 ± 0.05 | ND | ND | 11.83±2.74^a^ |
|  | LIF6 | 1.12 ± 0.66^a^ | 0.36 ± 0.02^a^ | 0.16 ± 0.04^a^ | ND | 0.20 ± 0.08 | ND | ND | ND | 1.85±0.60^a^ |
|  | LIF7 | 10.00 ± 2.03^a,b,c^ | 11.50 ± 2.32^a,b,c,d^ | 0.91 ± 0.13^a^ | ND | 0.60 ± 0.07 | ND | 0.33 ± 0.09^a,b^ | 0.21 ± 0.02^a^ | 23.56±4.63^a^ |
|  | LIF8 | 6.16 ± 1.24^a,b,c^ | 12.67 ± 1.38^a,b,c,d^ | ND | ND | 2.95 ± 0.06 | ND | ND | ND | 21.79±0.60^b^ |
|  | LIF9 | 14.64 ± 0.14^c^ | 18.14 ± 0.85^c,d^ | 1.37 ± 0.24^a^ | 0.26 ± 0.37 | 1.53 ± 0.11 | 0.22 ± 0.01 | 0.27 ± 0.05^a,b^ | 0.12 ± 0.03^a^ | 36.55±0.75^a^ |
|  | LIF10 | 8.06 ± 0.46^a,b,c^ | 10.40 ± 0.59^a,b,c,d^ | 0.73 ± 0.06^a^ | ND | 1.02 ± 0.7 | 0.77 ± 0.88 | 0.37 ± 0.26^a,b^ | 0.16 ± 0.08^a^ | 21.53±0.60^a^ |
|  | LIF11 | 0.83 ± 0.04^a^ | 0.55 ± 0.24^a^ | 0.22 ± 0.00^a^ | 8.66 ± 12.24 | 0.16 ± 0.13 | ND | 0.09 ± 0.07^a,b^ | 0.09 ± 0.05^a^ | 10.61±2.43^a^ |
|  | LIF12 | 0.59 ± 0.27^a^ | 3.72 ± 3.90^a,b,c^ | 0.59 ± 0.54^a^ | 1.81 ± 1.86 | 1.31 ± 1.62 | 0.89 ± 0.89 | 0.16 ± 0.10^a,b^ | 0.15 ± 0.02^a^ | 9.22±6.19^a^ |
|  | LIF13 | 7.52 ± 2.12^a,b,c^ | 10.61 ± 0.64^a,b,c,d^ | 1.85 ± 0.81^a^ | 0.43 ± 0.61 | 0.53 ± 0.61 | 0.31 ± 0.01 | 0.36 ± 0.25^a,b^ | 0.19 ± 0.26^a^ | 21.80±5.14^a^ |
|  | LIF14 | 14.00 ± 3.10^b,c^ | 20.49 ± 0.75^d^ | 1.69 ± 0.30^a^ | ND | 1.42 ± 2.01 | 0.09 ± 0.12 | 0.65 ± 0.40^a,b,c^ | 0.28 ± 0.27^a,b^ | 38.62±5.02^ab^ |
|  | LIF15 | 35.71 ± 13.23^d^ | 44.81 ± 18.23^e^ | 11.15 ± 8.10^b^ | 1.26 ± 0.38 | 2.77 ± 1.37 | 0.71 ± 0.00 | 1.25 ± 0.35^c^ | 0.38 ± 0.14^a,b^ | 98.06±31.81^a^ |

Values reported as μg per scoop (9 grams) of powder or per liquid equivalent to 1 ready to feed bottle( 60 mL). Identical letters within columns indicate that samples are not statistically different, according to analysis of variance and Tukey’s mean comparison test (p < 0.05).

**Table S4.** Tocopherols profile of liquid and powder IF.

| **Type** | **Sample** | **α-Tocopherol** | **β-Tocopherol** | **γ-Tocopherol** | **δ-Tocopherol** | **Total Tocopherols** |
| --- | --- | --- | --- | --- | --- | --- |
| Powder | PIF1 | 0.081 ± 0.028 | ND | 0.029 ± 0.019 | 0.062 ± 0.007^a^ | 0.172 ± 0.040^a,b^ |
|  | PIF2 | 0.014 ± 0.017 | ND | 0.020 ± 0.001 | 0.120 ± 0.050^a^ | 0.155 ± 0.077^a,b^ |
|  | PIF3 | ND | 0.018 ± 0.008^a,b^ | 0.064 ± 0.037 | 0.451 ± 0.203^a,b^ | 0.532 ± 0.248^b,c^ |
|  | PIF4 | 0.016 ± 0.016 | 0.004 ± 0.001^a,b^ | 0.029 ± 0.023 | 0.276 ± 0.104^a,b^ | 0.325 ± 0.144^a,b,c^ |
|  | PIF5 | 0.055 ± 0.039 | 0.029 ± 0.030^a,b^ | 0.062 ± 0.050 | 0.278 ± 0.030^a,b^ | 0.424 ± 0.049^a,b,c^ |
|  | PIF6 | 0.030 ± 0.025 | 0.044 ± 0.051^a,b^ | 0.056 ± 0.068 | 0.613 ± 0.252^b^ | 0.743 ± 0.397^c^ |
|  | PIF7 | 0.017 ± 0.006 | 0.020 ± 0.005^a,b^ | 0.028 ± 0.017 | 0.171 ± 0.045^a,b^ | 0.237 ± 0.060^a,b^ |
|  | PIF8 | 0.036 ± 0.043 | 0.010 ± 0.010^a,b^ | 0.022 ± 0.021 | 0.098 ± 0.035^a^ | 0.166 ± 0.109^a,b^ |
|  | PIF9 | 0.015 ± 0.012 | 0.012 ± 0.010^a,b^ | 0.017 ± 0.006 | 0.027 ± 0.015^a^ | 0.071 ± 0.030^a,b^ |
|  | PIF10 | 0.025 ± 0.003 | 0.010 ± 0.003^a,b^ | 0.009 ± 0.001 | 0.110 ± 0.000^a^ | 0.155 ± 0.005^a,b^ |
|  | PIF11 | 0.037 ± 0.006 | 0.015 ± 0.006^a,b^ | 0.034 ± 0.004 | 0.294 ± 0.012^a,b^ | 0.380 ± 0.016^a,b,c^ |
|  | PIF12 | 0.021 ± 0.016 | 0.023 ± 0.009^a,b^ | 0.059 ± 0.020 | 0.262 ± 0.039^a,b^ | 0.364 ± 0.034^a,b,c^ |
|  | PIF13 | 0.026 ± 0.033 | 0.006 ± 0.005^a,b^ | 0.021 ± 0.018 | 0.101 ± 0.042^a^ | 0.154 ± 0.098^a,b^ |
|  | PIF14 | 0.046 ± 0.013 | 0.010 ± 0.001^a,b^ | 0.021 ± 0.007 | 0.116 ± 0.033^a^ | 0.193 ± 0.041^a,b^ |
|  | PIF15 | 0.058 ± 0.054 | 0.012 ± 0.005^a,b^ | 0.017 ± 0.002 | 0.082 ± 0.008^a^ | 0.169 ± 0.043^a,b^ |
| Liquid | LIF1 | 0.017 ± 0.006 | 0.027 ± 0.027^a,b^ | 0.014 ± 0.016 | 0.047 ± 0.022^a^ | 0.106 ± 0.028^a,b^ |
|  | LIF2 | 0.042 ± 0.048 | 0.006 ± 0.001^a,b^ | 0.000 ± 0.000 | 0.019 ± 0.003^a^ | 0.067 ± 0.050^a^ |
|  | LIF3 | 0.020 ± 0.003 | 0.004 ± 0.006^a,b^ | 0.000 ± 0.000 | 0.030 ± 0.042^a^ | 0.054 ± 0.051^a^ |
|  | LIF4 | 0.054 ± 0.076 | 0.097 ± 0.016^a,b^ | 0.000 ± 0.000 | 0.177 ± 0.003^a,b^ | 0.328 ± 0.096^a,b,c^ |
|  | LIF5 | 0.049 ± 0.013 | 0.048 ± 0.025^a,b^ | 0.051 ± 0.038 | 0.188 ± 0.011^a,b^ | 0.335 ± 0.036^a,b,c^ |
|  | LIF6 | 0.092 ± 0.043 | 0.055 ± 0.022^a,b^ | ND | 0.123 ± 0.045^a^ | 0.270 ± 0.110^a,b^ |
|  | LIF7 | 0.108 ± 0.081 | 0.040 ± 0.029^a,b^ | ND | 0.083 ± 0.062^a^ | 0.231 ± 0.172^a,b^ |
|  | LIF8 | 0.116 ± 0.020 | 0.035 ± 0.030^a,b^ | ND | 0.085 ± 0.023^a^ | 0.235 ± 0.034^a,b^ |
|  | LIF9 | 0.011 ± 0.011 | 0.000 ± 0.000^a^ | ND | 0.047 ± 0.002^a^ | 0.058 ± 0.013^a^ |
|  | LIF10 | 0.037 ± 0.009 | 0.006 ± 0.004^a,b^ | ND | 0.036 ± 0.004^a^ | 0.079 ± 0.017^a,b^ |
|  | LIF11 | 0.008 ± 0.003 | 0.006 ± 0.002^a,b^ | ND | 0.055 ± 0.004^a^ | 0.069 ± 0.009^a,b^ |
|  | LIF12 | 0.009 ± 0.005 | 0.009 ± 0.000^a,b^ | ND | 0.041 ± 0.011^a^ | 0.058 ± 0.015^a^ |
|  | LIF13 | 0.036 ± 0.022 | 0.099 ± 0.094^b^ | ND | 0.087 ± 0.069^a^ | 0.222 ± 0.142^a,b^ |
|  | LIF14 | 0.089 ± 0.021 | ND | ND | 0.144 ± 0.102^a^ | 0.233 ± 0.123^a,b^ |
|  | LIF15 | 0.094 ± 0.056 | ND | ND | 0.049 ± 0.000^a^ | 0.143 ± 0.056^a,b^ |

Values reported in mg per scoop (9 grams) of powder or per liquid equivalent to1 ready to feed bottle, (60 mL). Identical letters within columns indicate that samples are not statistically different, according to analysis of variance and Tukey’s mean comparison test (p < 0.05).
